# Safety and Immunogenicity of Omicron BA.4/BA.5 Bivalent Vaccine Against Covid-19

**DOI:** 10.1101/2022.12.11.22283166

**Authors:** Spyros Chalkias, Jordan Whatley, Frank Eder, Brandon Essink, Shishir Khetan, Paul Bradley, Adam Brosz, Nichole McGhee, Joanne E. Tomassini, Xing Chen, Xiaoping Zhao, Andrea Sutherland, Xiaoying Shen, Bethany Girard, Darin K. Edwards, Jing Feng, Honghong Zhou, Stephen Walsh, David C. Montefiori, Lindsey R. Baden, Jacqueline M. Miller, Rituparna Das

**Author notes:** **Corresponding Author:** Spyros Chalkias, M.D., Moderna, Inc., Cambridge, MA, USA 02141-2025.

## Abstract

**Background:** Information on the safety and immunogenicity of the omicron BA.4/BA.5-containing bivalent booster mRNA-1273.222 are needed.

**Methods:** In this ongoing, phase 2/3 trial, 50-μg mRNA-1273.222 (25-μg each ancestral Wuhan-Hu-1 and omicron BA.4/BA.5 spike mRNAs) is compared to 50-μg mRNA-1273, administered as second boosters in adults who previously received a 2-injection (100-μg) primary series and first booster (50-μg) dose of mRNA-1273. The primary objectives were safety and immunogenicity 28 days post-boost.

**Results:** Participants received 50-μg of mRNA-1273 (n=376) or mRNA-1273.222 (n=511) as second booster doses. Omicron BA.4/BA.5 and ancestral SARS-CoV-2 D614G neutralizing antibody geometric mean titers (GMTs [95% confidence interval]) after mRNA-1273.222 (2324.6 [1921.2-2812.7] and 7322.4 [6386.2-8395.7]) were significantly higher than mRNA-1273 (488.5 [427.4-558.4] and 5651.4 (5055.7-6317.3) respectively, at day 29 post-boost in participants with no prior SARS-CoV-2-infection. A randomly selected subgroup (N=60) of participants in the mRNA-1273.222 group also exhibited cross-neutralization against the emerging omicron variants BQ.1.1 and XBB.1. No new safety concerns were identified with mRNA-1273.222.

Vaccine effectiveness was not assessed in this study; in an exploratory analysis 1.6% (8/511) of mRNA-1273.222 recipients had Covid-19 post-boost.

**Conclusion:** The bivalent omicron BA.4/BA.5-containing vaccine mRNA-1273.222 elicited superior neutralizing antibody responses against BA.4/BA.5 compared to mRNA-1273, with no safety concerns identified.

(Supported by Moderna; ClinicalTrials.gov Identifier: NCT04927065)

## INTRODUCTION

Booster immunization against severe acute respiratory syndrome coronavirus 2 (SARS-CoV-2) improves immune responses and vaccine effectiveness against coronavirus disease 2019 (Covid-19).^1-8^ The emergence of antigenically-divergent omicron variants required variant-targeting immunization strategies and we previously evaluated the safety and immunogenicity of an omicron BA.1-containing bivalent booster (mRNA-1273.214) that has been deployed in multiple geographies.^5,9-12^ The BA.1 bivalent vaccine was well-tolerated and increased the magnitude and the durability of the neutralizing antibody responses against BA.1 compared to the original booster mRNA-1273.^5,6^ Additionally, mRNA-1273.214 exhibited cross-neutralization ability against omicron BA.4/BA.5, BA.2.75 and other variants.^5,6^

An omicron BA.4/BA.5 bivalent booster (mRNA-1273.222) has been authorized in the United States and elsewhere for the fall 2022 immunization campaign.^13^ Early effectiveness data demonstrate that recipients of an omicron-containing bivalent booster have improved protection against Covid-19 relative to immunization with the original mRNA-1273 vaccine only.^13-15^ However, clinical information on the safety and immunogenicity of the omicron BA.4/BA.5 bivalent booster has not been previously presented. In addition, following the BA.5 infection waves in the summer and early fall of 2022, other omicron variants with additional spike protein mutations that are likely to confer immune escape have emerged. Specifically, omicron BQ.1.1, a sublineage of BA.5 and omicron XBB.1, a recombinant of BA.2.10.1 and BA.2.75 sublineages are being monitored globally.^16^ Herein, we summarize mRNA-1273.222 interim safety and immunogenicity results from an ongoing phase 2/3 clinical trial and we also assess the neutralization activity for both omicron bivalent booster vaccines, mRNA-1273.222 and mRNA-1273.214 against emerging divergent variants.

## METHODS

### Trial Oversight and Participants

This open-label, ongoing phase 2/3 study (NCT04927065) evaluates the safety, reactogenicity and immunogenicity of the bivalent omicron BA.4/BA.5-containing mRNA-1273.222 booster vaccine compared to the currently-authorized mRNA-1273 booster in adults who had previously received a 2-injection primary series (100-μg) and first booster doses (50-μg) of mRNA-1273 in Coronavirus Efficacy (COVE) trial^17,18^ or under US emergency use authorization (EUA), enrolled in a sequential, non-randomized manner. Participants received single second boosters of 50-μg mRNA-1273 (part F, cohort 2, enrolled between February 18^th^ and March 8^th^, 2022) or 50-μg bivalent mRNA-1273.222 (part H, enrolled between August 10^th^ and 23^rd^, 2022). The non-contemporaneous mRNA-1273 group serves as a within-study comparator for the pre-specified immunogenicity comparison between mRNA-1273.222 and mRNA-1273 given that enrollment with the original vaccine mRNA-1273 was no longer feasible after the authorization of bivalent booster vaccines.

The trial is being conducted across 23 US sites, in accordance with the International Council for Harmonisation of Technical Requirements for Registration of Pharmaceuticals for Human Use, Good Clinical Practice guidelines. The central Institutional Review Board approved the protocol and consent forms. All participants provided written informed consent.

Adults with a known history of SARS-CoV-2 infection ≤3 months from screening were excluded. Additional details of inclusion/exclusion criteria, trial oversight and study design are provided in the Supplemental Material.

### Trial vaccine

Bivalent mRNA-1273.222 50-μg vaccine contains two mRNAs (25-μg each) encoding the prefusion-stabilized spike glycoproteins of ancestral SARS-CoV-2 (Wuhan-Hu-1) and the omicron variant (BA.4/BA.5). The monovalent mRNA-1273 50-μg original vaccine contains a single mRNA encoding the spike glycoprotein of ancestral SARS-CoV-2 (Wuhan-Hu-1). mRNA-1273.222 and mRNA-1273 were administered intramuscularly at 50-μg in a 0.5 mL volume.

### Safety Assessment

The primary safety objective was to evaluate the safety and reactogenicity of 50-μg mRNA-1273.222 when administered as a second booster dose. Safety assessments included solicited local and systemic adverse reactions ≤7 days and unsolicited adverse events ≤28 days post-booster administration, and serious adverse events, adverse events leading to discontinuation from study vaccine and/or participation, medically-attended adverse events, and adverse events of special interest from day 1 through the entire study period (∼6 months).

### Immunogenicity assessment

The pre-specified primary immunogenicity objectives were to demonstrate non-inferior nAb responses (based on geometric mean titer [GMT] ratio [GMR] and seroresponse rate [SRR] difference) or superior nAb responses (based on GMR) against omicron BA.4/BA.5, and non-inferior nAb responses (based on GMR and SRR difference) against ancestral SARS-CoV-2 with the D614G mutation (ancestral SARS-CoV-2 [D614G]), 28 days after the second booster dose (day 29) of mRNA 1273.222 50-μg compared with mRNA-1273 50-μg (Table S1 and statistical methods).

Neutralizing antibody GMTs at inhibitory dilutions 50% (ID50) were assessed using validated SARS-CoV-2 spike-pseudotyped lentivirus neutralization assays against pseudoviruses containing the SARS-CoV-2 full-length spike proteins of ancestral SARS-CoV-2 (D614G), or omicron variants BA.4/BA.5, BQ.1.1 and XBB.1 variants. Geometric mean (GM)-levels of spike-binding antibody (bAb) were also assessed using an (Meso Scale Discovery [MSD]) assay against ancestral SARS-CoV-2 (D614G), gamma (P.1), alpha (B.1.1.7), delta [B.1.617.2; AY.4], and omicron (BA.1) variants. These immunogenicity assays are further described in the supplement.

### Incidence of SARS-CoV-2 Infections

The incidence of symptomatic and asymptomatic SARS-CoV-2 infection was an exploratory objective (Table S1 and Supplementary Methods).

### Statistical analysis

Statistical analysis methods are detailed in the Supplemental Appendix and analysis sets in Table S2 and Fig. S1. Safety was evaluated in the safety set consisting of all participants who received mRNA-1273.222. Solicited ARs were assessed in all participants and by pre-booster SARS-CoV-2 infection status in the solicited safety set. The per-protocol immunogenicity set consists of all participants who received the planned booster doses, had pre-booster and day 29 antibody data available and no major protocol deviations. Primary immunogenicity objectives were assessed in the per-protocol immunogenicity–SARS-CoV-2-negative set (primary analysis set). Analyses were also performed in participants who had evidence of SARS-CoV-2-infection pre-booster.

The primary immunogenicity objectives were evaluated using a pre-specified hierarchical approach (Fig. S2 and Supplemental Methods). An interim analysis was planned at day 29 with a two-sided alpha (0.05) for the immunogenicity hypothesis testing. The superiority of the antibody response against omicron BA.4/BA.5 after a second booster dose of 50-μg mRNA-1273.222 compared with 50-μg mRNA-1273 was evaluated only after meeting non-inferiority criteria for the four primary objectives:^19^ non-inferiority of the antibody response against BA.4/BA.5 after the second booster doses of 50-μg mRNA-1273.222 versus 50-μg mRNA-1273 based on GMR (1) and SRR difference (2), and non-inferiority of the antibody response against ancestral SARS-CoV-2 (D614G) after the second booster doses of 50-μg mRNA-1273.222 versus 50-μg mRNA-1273 based on GMR (3) and SRR difference (4); all statistical testing was based on alpha of 0.05 (two-sided) at day 29. Non-inferiority was considered met when the lower bound of the 95.0% confidence interval (CI) of GMR is >0.667 and the seroresponse rate-difference for BA.4/BA.5 is >-5% and seroresponse rate-difference for ancestral SARS-CoV-2 (D614G) is >-10%. Superiority is considered met when the lower bound of the 95.0% CI of GMR is >1.^20,21^

Observed GMTs (95% CI) using t-distribution of log-transformed antibody titers are presented. Differences in antibody responses between the mRNA-1273.222 and mRNA-1273 groups were assessed using an analysis of covariance (ANCOVA) model (antibody titers post-booster as dependent variable, group variable mRNA-1273.222 and mRNA-1273 as the fixed effect) adjusted for age groups (<65, ≥65 years) and pre-booster antibody titers. The GMTs (95% CI) estimated by the geometric least square mean from the model and differences in antibody responses (GMR) between groups estimated by the ratio of geometric least mean square (95.0% CIs) are provided. Seroresponses (change from <lower limit of quantification [LLOQ] to ≥4 × LLOQ, or at least a 4-fold rise if baseline is ≥LLOQ) with 95% CI (Clopper-Pearson) and SRR differences between mRNA-1273.222 and mRNA-1273 groups (95.0% CI; stratified Miettinen-Nurminen) adjusting for age groups are provided.

Additional analyses described in the Supplementary Methods included assessment of the primary immunogenicity endpoints for participants with previous SARS-CoV-2-infection and observed bAb GM-levels against SARS-CoV-2 variants and bAb comparisons between mRNA-1273.222 and mRNA-1273 based on ANCOVA-estimated antibody levels and GMRs (95% CIs). An analysis of observed GMTs against the omicron BQ.1.1 and XBB.1 was also performed in random subsets of recipients (n=60; 40 without prior infection and 20 with prior infection) in the mRNA-1273.222 group and the previously described omicron BA.1-containing bivalent booster mRNA-1273.214 group^5^ (supplementary Table S3). Additionally, whether time intervals (between the first booster dose of mRNA-1273 and second booster doses of mRNA-1273 and mRNA-1273.222) affected the neutralizing antibody response post-boost were explored in participants without previous infection grouped by quartiles of dosing intervals within each booster vaccine group, and omicron BA.4/BA.5 GMT and GMFR at day 29 were summarized by interval groups. The number and percentage of participants with asymptomatic or symptomatic SARS-CoV-2-infection and Covid-19-events post-booster are summarized. All analyses were conducted using SAS Version 9.4 or higher.

## RESULTS

### Trial population

Between August 10 and 23 2022, 511 participants received 50-μg of mRNA-1273.222. These participants had previously received the primary series of 100-μg mRNA-1273 and a first booster dose of 50-μg mRNA-1273, ≥3 months before enrollment (Fig. 1). Of these, 305 (59.7%) participants originated from the COVE trial and 206 (40.3%) were US vaccinees under the EUA, respectively. Given that within the study enrollment windows the original booster vaccine mRNA-1273 was no longer authorized, a mRNA-1273 comparator group was not enrolled. Instead, a within-study, non-contemporaneous mRNA-1273 group (n=379, enrollment February 18th-March 8th, 2022)^5^ was used for the immunogenicity comparison with 264 (69.7%) COVE participants and 115 (30.3%) US EUA vaccinees, respectively. Four (0.8%) participants in the mRNA-1273.222 and 6 (1.5%) in the mRNA-1273 groups discontinued the study.

**Figure 1.**
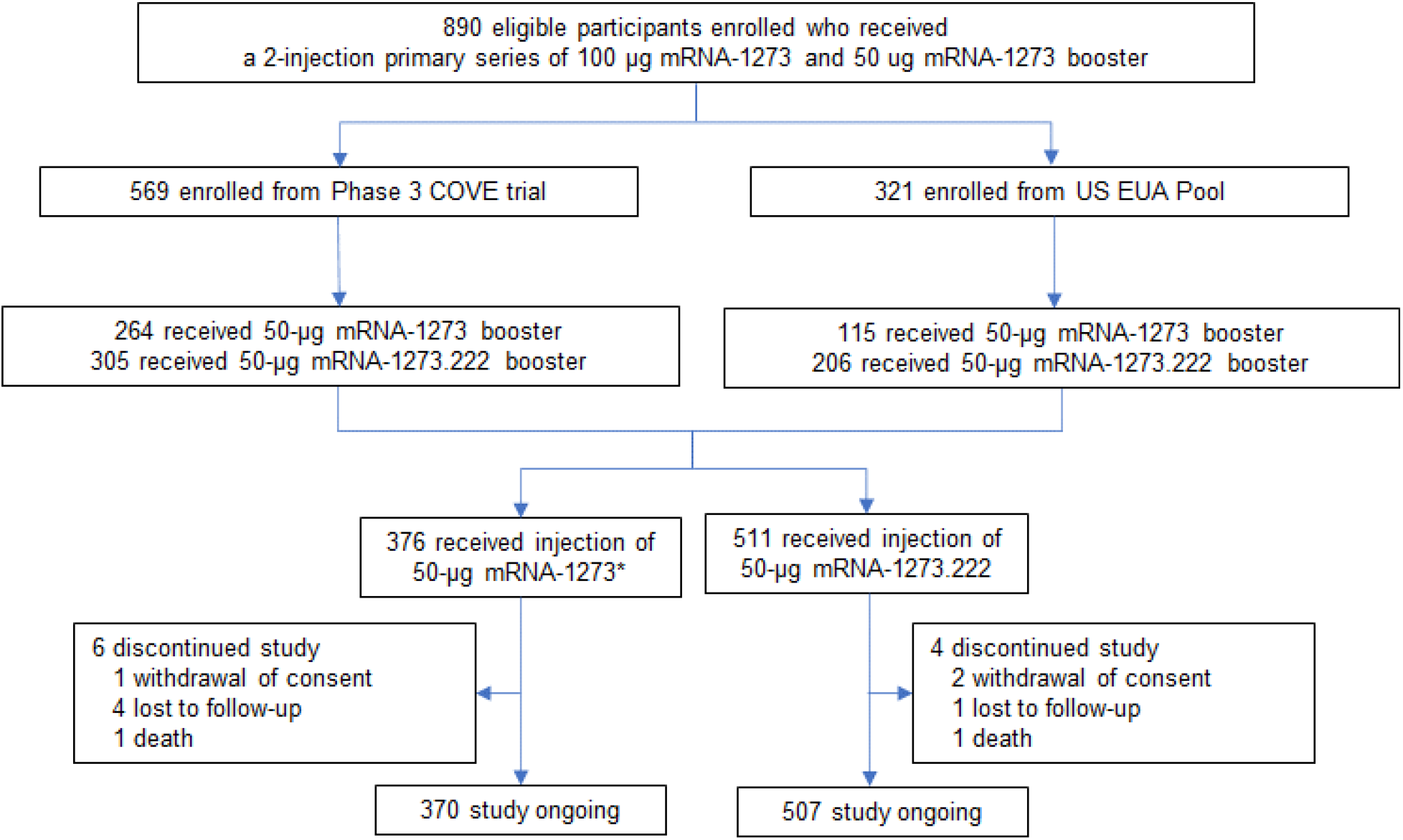
Trial Profile. Emergency Use Authorization, EUA; Coronavirus Efficacy, COVE. Eligible participants who received a prior 2-injection primary series of 100-μg mRNA-1273 and a 50-μg mRNA-1273 booster dose either in the COVE trial or under the US EUA were enrolled to receive a second booster dose of 50-μg mRNA-1273 or mRNA-1273.222. *379 participants were enrolled and received mRNA-1273; two participants had previously received the primary series but not a first booster dose and another participant had a major protocol deviation, and three were excluded from all analysis sets. Data cutoff date was September 23, 2022. The within-study non-contemporaneous mRNA-1273 group (Part F, cohort 2) was enrolled during February 18th-March 8th, 2022 and data for this group is based on the data cutoff date of July 6, 2022 at the day 91 interim analysis.

Participant demographics and baseline characteristics are shown in Table 1. Mean ages were 50.8 and 57.6 years, and 62% and 51% were female in the 50-μg mRNA-1273.222 and 50-μg mRNA-1273 groups, respectively. Most participants were White (83% and 86%) and 11% and 10% were of Hispanic/Latinx ethnicity in the mRNA-1273.222 and mRNA-1273 groups, respectively. The percentages of participants with SARS-CoV-2-infection pre-booster were 56% in the mRNA-1273.222 and 27% in the mRNA-1273 groups. Median time (days [IQR)]) were similar between second doses of mRNA-1273 in the primary series and the first booster of mRNA-1273 (251 [228-299] and 242 [225-260]) and the second booster doses (289 [258-312] in the mRNA-1273.222 group than mRNA-1273 (134 [118-150]) group.

**Table 1.** Demographics and Study Participant Characteristics, Safety Set.

| Characteristics n (%)* | mRNA-1273.222 50 µg<br>N=511 | mRNA-1273 50 µg<br>N=376 |
| --- | --- | --- |
| Age at Screening (yr) |  |  |
| Mean (range) | 50.8 (19-89) | 57.6 (20-96) |
| Age subgroup |  |  |
| ≥18 and <65 years | 406 (79.5) | 226 (60.1) |
| ≥65 years | 105 (20.5) | 150 (39.9) |
| Gender |  |  |
| Male | 195 (38.2) | 186 (49.5) |
| Female | 316 (61.8) | 190 (50.5) |
| Ethnicity |  |  |
| Hispanic or Latino | 58 (11.4) | 37 (9.8) |
| Not Hispanic or Latino | 448 (87.7) | 339 (90.2) |
| Not reported or unknown | 5 (1.0) | 0 |
| Race |  |  |
| White | 426 (83.4) | 322 (85.6) |
| Black or African American | 56 (11.0) | 28 (7.4) |
| Asian | 11 (2.2) | 16 (4.3) |
| American Indian or Alaska Native | 1 (0.2) | 1 (0.3) |
| Native Hawaiian or Other Pacific Islander | 0 | 1 (0.3) |
| Multiracial | 8 (1.6) | 2 (0.5) |
| Other | 6 (1.2) | 2 (0.5) |
| Not reported or unknown | 3 (0.6) | 4 (1.1) |
| Time between second injection of mRNA-1273 in the primary series and the first booster of mRNA-1273 (days) |  |  |
| n | 509 | 374 |
| Median | 251.0 | 242.0 |
| IQR | (228-299) | (225-260) |
| Time between first booster dose of mRNA-1273 and the second booster (days) |  |  |
| n | 509 | 374 |
| Median | 289.0 | 134.0 |
| IQR | (258-312) | (118-150) |
| Pre-booster RT-PCR SARS-CoV-2 |  |  |
| Negative | 488 (95.5) | 366 (97.3) |
| Positive | 10 (2.0) | 2 (0.5) |
| Missing | 13 (2.5) | 8 (2.1) |
| Pre-booster antibody to SARS-CoV-2 nucleocapsid† |  |  |
| Negative | 226 (44.2) | 276 (73.4) |
| Positive | 282 (55.2) | 100 (26.6) |
| Missing | 3 (0.6) | 0 |
| Pre-booster SARS-CoV-2 status‡ |  |  |
| Negative | 216 (42.3) | 267 (71.0) |
| Positive | 286 (56.0) | 101 (26.9) |
| Positive by both RT-PCR and SARS-CoV-2 nucleocapsid† | 6 (1.2) | 1 (0.3) |
| Positive by RT-PCR only | 4 (0.8) | 1 (0.3) |
| Positive by SARS-CoV-2 nucleocapsid only† | 276 (54.0) | 99 (26.3) |
| Missing | 9 (1.8) | 8 (2.1) |
| IQR=interquartile range; RT-PCR = reverse transcription polymerase chain reaction. *Percentages based on the number of participants in the safety set. †Elecsys assay for binding antibody to SARS-CoV-2 nucleocapsid. ‡Pre-booster SARS-CoV-2 status was positive if there was evidence of prior SARS-CoV-2-infection, defined as positive binding antibody against the SARS-CoV-2 nucleocapsid or positive RT-PCR at day 1; Negative SARS-CoV-2 status was defined as negative binding antibody against the SARS-CoV-2 nucleocapsid and a negative RT-PCR at day 1. |  |  |

### Safety

The safety of the mRNA-1273 group has been previously presented ^5^. Median duration of follow-up days (IQR) was 37.0 (33.0-39.0) for the mRNA-1273.222 participants and the most frequently reported local adverse reaction after administration of mRNA-1273.222 was injection-site pain. The most frequent systemic reactions were fatigue, headache, myalgia, and arthralgia (Fig. 2 and Table S4). The majority of solicited adverse reactions were mild-to-moderate (grades 1-2) and the most common grade 3 reactions were fatigue and myalgia; no grade 4 reactions occurred. The frequency of local and systemic adverse reactions was 84.7% and 77.3%, respectively in participants without prior SARS-CoV-2 infection and 81.2% and 69.6% in those with prior SARS-CoV-2 infection.

**Figure 2.**
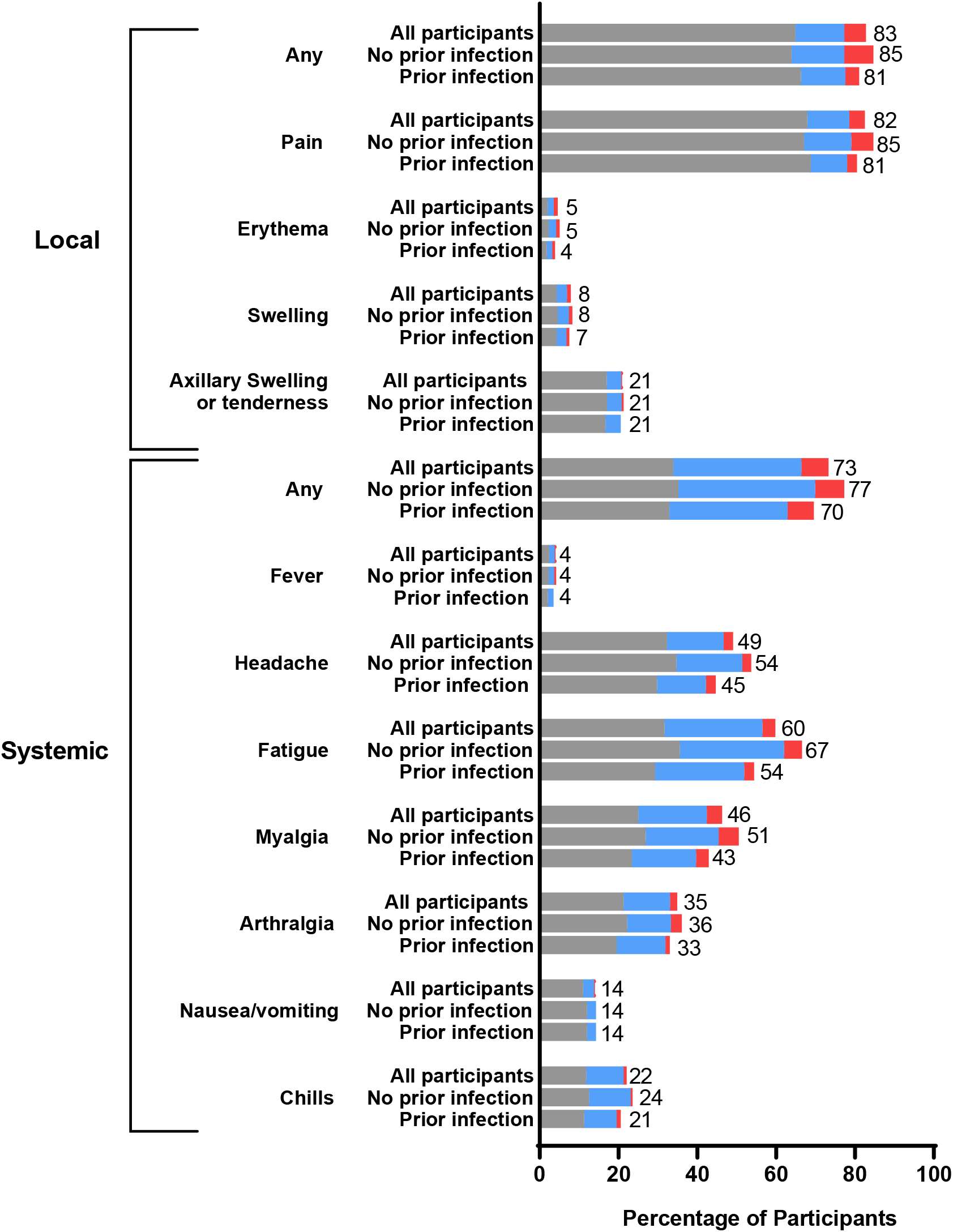
Solicited Local and Systemic Adverse Reactions. Percentages of participants who had solicited local or systemic adverse reactions within 7 days by grade after the 50 μg mRNA-1273.222 second booster dose in all participants and those with and without SARS-CoV-2 infection at pre-booster in the solicited safety set.

The incidence of unsolicited adverse events regardless of the relationship to vaccination ≤28 days after the booster dose of mRNA-1273.222 was 23% (Table S5)^6^ and the incidence of those considered related to study vaccination by the investigator was 8%. Three (0.6%) participants (60-70 years of age) in the mRNA-1273.222 group experienced serious adverse events; none were considered related to study vaccination by the investigators. These included events of anginal equivalent and syncope and anemia in two participants with medical histories of vascular and cardiac disease and anemia and hepatic cirrhosis, respectively, and one fatal event 7 days post-immunization of subarachnoid hemorrhage in a participant with vascular risk factors. Medically-attended adverse events occurred in 14% of mRNA-1273.222 participants; none were considered related to study vaccination by investigators. Two grade 3 events of fatigue, considered related to study vaccination, were reported. No events of myocarditis or pericarditis and no adverse events leading to study discontinuation occurred in this interim analysis.

### Immunogenicity

In participants without evidence of prior SARS-CoV-2 28 days after the mRNA-1273.222 and mRNA-1273 booster dose, respectively, the observed nAb GMTs (95% CI) were 2324.6 (1921.2−2812.7) and 488.5 (427.4−558.4) against omicron BA.4/BA.5 and were 7322.4 (6386.2−8395.7) and 5651.4 (5055.7−6317.3) against ancestral SARS-CoV-2 (D614G) (Figs. 3 and S3 and Table 2). ANCOVA-estimated GMTs after adjusting for age groups and pre-booster titers 28 days after the mRNA-1273.222 and mRNA-1273 booster dose, respectively, were 2747.3 (2399.2−3145.9) and 436.7 (389.1−490.0) against omicron BA.4/BA.5 and were 9555.8 (8593.6−10625.7) and 4882.2 (4457.7−5347.1) against ancestral SARS-CoV-2 (D614G). The GMR (95.0% CI) against omicron BA/BA.5 was 6.29 (5.27−7.51) meeting the pre-specified superiority criterion. The GMR (95.0% CI) against ancestral SARS-CoV-2 (D614G) was 1.96 (1.70−2.25), meeting the pre-specified criterion for non-inferiority. Seroresponse rates (95% CI) against omicron BA.4/BA.5 were 98.1% (95.2−99.5%) for mRNA-1273.222 and 86.4% (81.6−90.3%) for mRNA-1273, and seroresponse rates against ancestral SARS-CoV-2 (D614G) were 100% (98.3−100% and 98.6−100%) 28 days after the mRNA-1273.222 and mRNA-1273 booster doses, respectively, with estimated differences of 12.1% (6.9−17.3%) and 0, both meeting the pre-specified non-inferiority criterion. Therefore, all primary and immunogenicity endpoints were met (Fig. S2). Neutralizing titers against omicron BA.4/BA.5 and ancestral SARS-CoV-2 (D614G) were also consistently higher with mRNA-1273.222 than mRNA-1273 at day 29 among those ≥65 years and 18-<65 years of age (Fig. S4). In participants with evidence of prior SARS-CoV-2-infection, GMTs were higher following the mRNA-1273.222 than mRNA-1273 booster against both omicron BA.4/BA.5 and ancestral SARS-CoV-2 (D614G) with GMRs (95% CI) of 5.11 (4.10−6.36) and 1.84 (1.56−2.18), respectively (Fig. 3; Table S6).

**Figure 3:**
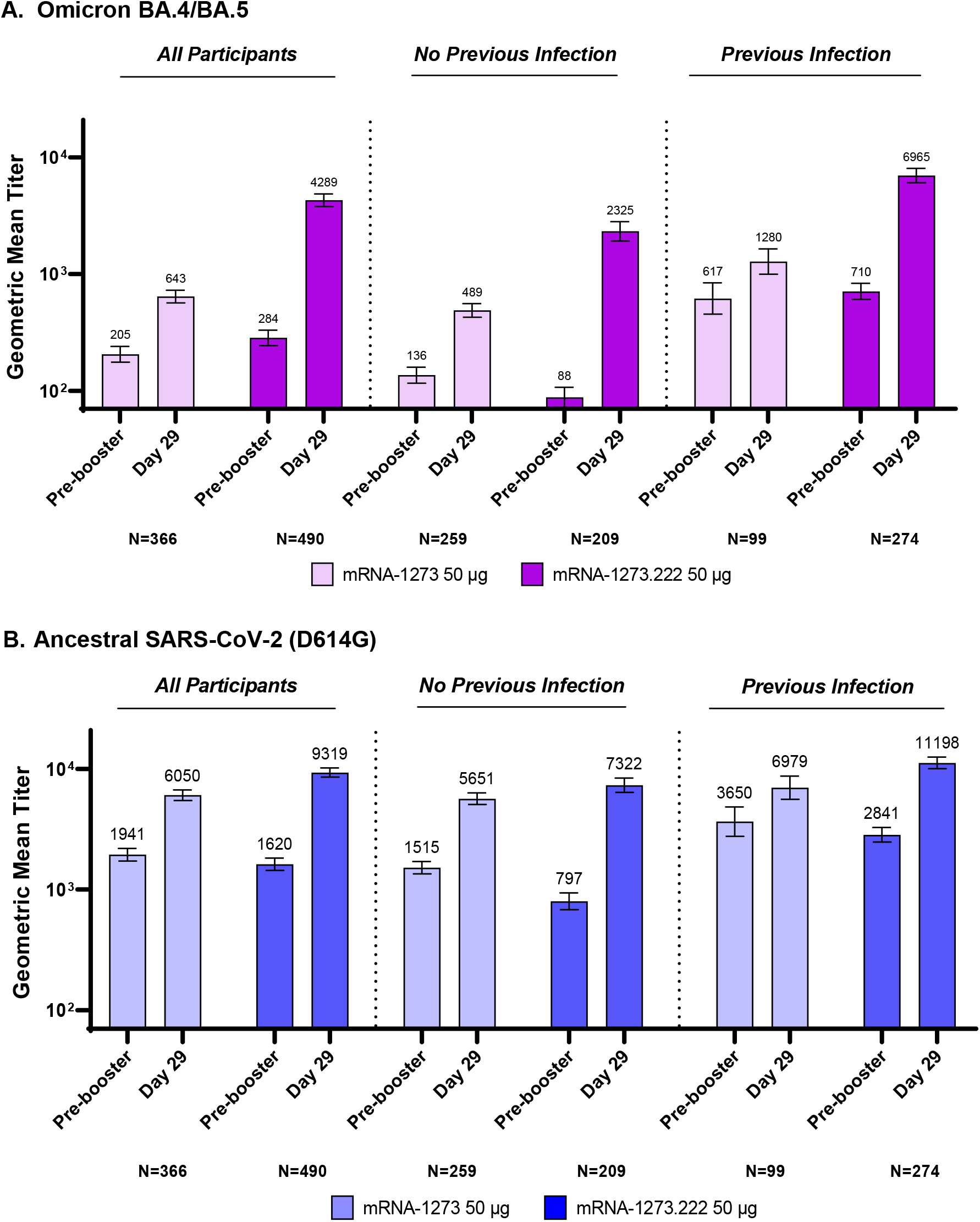
Observed Neutralizing Antibody Titers Against Ancestral SARS-CoV-2 (D614G) and Omicron BA.4/BA.5 after 50-μg of mRNA-1273.222 and mRNA-1273 Administered as Second Booster Doses. GM = geometric mean; CI = confidence interval. Pseudovirus neutralizing antibody geometric mean titers are provided for all participants regardless of SARS-CoV-2-infection pre-booster (per-protocol immunogenicity set), and those with and without prior SARS-CoV-2-infection pre-booster. Data are from participants with non-missing data at the timepoint. Seven participants in mRNA-1273.222 and eight participants in the mRNA-1273 group were missing pre-booster SARS-CoV-2 status. Antibody values reported as below the lower limit of quantification (LLOQ: 18.5 for ancestral SARS-CoV-2 (D614G); 36.7 for omicron BA.4/BA.5) were replaced by 0.5 x LLOQ. Values greater than the upper limit of quantification (ULOQ: 45,118 for ancestral SARS-CoV-2 (D614G); 13,705 for omicron BA.4/BA.5) were replaced by the ULOQ if actual values are not available. 95% CIs were calculated based on the t-distribution of the log-transformed values for GM value, then back transformed to the original scale for presentation. Data for observed nAb GMTs by prior SARS-CoV-2-infection are provided in Table S8.

**Table 2.** Primary Immunogenicity Analysis of Omicron BA4/BA.5 and Ancestral SARS-CoV-2 (D614G) after 50-μg of mRNA-1273.222 and mRNA-1273 Administered as Second Booster Doses in Participants with No Prior SARS-CoV-2-Infection.

|  | Ancestral SARS-CoV-2 (D614G) |  | Omicron BA.4/BA.5 |  |
| --- | --- | --- | --- | --- |
|  | 50 µg<br>mRNA-1273.222<br>Booster Dose | 50 ug<br>mRNA-1273<br>Booster Dose | 50 µg<br>mRNA-1273.222<br>Booster Dose | 50 ug<br>mRNA-1273<br>Booster Dose |
|  | N=209 | N=259 | N=209 | N=259 |
| Pre-booster n† | 209 | 259 | 209 | 259 |
| Observed GMT (95% CI)§ | 796.9<br>(678.7-935.8) | 1515.4<br>(1347.5-1704.2) | 87.9<br>(72.2-107.1) | 136.1<br>(116.3-159.3) |
| Day 29, n† | 209 | 259 | 209 | 259 |
| Observed GMT (95% CI)§ | 7322.4<br>(6386.2-8395.7) | 5651.4<br>(5055.7-6317.3) | 2324.6<br>(1921.2-2812.7) | 488.5<br>(427.4-558.4) |
| GMFR (95% CI)§ | 9.2 (7.9-10.6) | 3.7 (3.4-4.1) | 26.4 (22.0-31.9) | 3.6 (3.3-4.0) |
| Estimated GMT (95% CI)¶¶ | 9555.8<br>(8593.6-10625.7) | 4882.2<br>(4457.7-5347.1) | 2747.3<br>(2399.2-3145.9) | 436.7<br>(389.1-490.0) |
| GMR (95% CI)¶¶\|\| | 1.96 (1.70- 2.25) |  | 6.29 (5.27-7.51) ¶¶\|\| |  |
| Day 29 SRR, injection 1,<br>n/N1, %‡ | 209/209, 100 | 259/259, 100 | 205/209, 98.1 | 222/257, 86.4 |
| (95% CI) | (98.3-100.0) | (98.6-100.0) | (95.2-99.5) | (81.6-90.3) |
| Difference (95% CI)\|\|†† | 0 |  | 12.1 (6.9-17.3) |  |
| Day 29 SRR, pre-booster,<br>n/N1, %‡ | 168/209, 80.4 | 111/259, 42.9 | 190/209, 90.9 | 98/259, 37.8 |
| (95% CI) | (74.3-85.5) | (36.7-49.1) | (86.2-94.4) | (31.9-44.0) |
| Difference (95% CI)\|\|†† | 37.3 (29.0-45.6) |  | 53.9 (46.7-61.2) |  |
| ANCOVA=analysis of covariance, CI=Confidence interval, GMT=geometric mean titer; GMFR=geometric mean fold-rise; GMR=Geometric mean ratio, mRNA-1273.222 vs mRNA-1273. ID50=50% inhibitory dose; LLOQ=lower limit of quantification; LS=least squares; SRR=seroresponse rate; ULOQ=upper limit of quantification. Antibody values assessed by pseudovirus neutralizing antibody assay reported as below the LLOQ (18.5 for ancestral [D614G] and 36.7 for omicron BA.4/BA.5) are replaced by 0.5 × LLOQ. Values greater than ULOQ (45,118 for ancestral [D614G] and 13,705 for omicron BA.4/BA.5) are replaced by the ULOQ if actual values are not available. Includes participants with no prior SARS-CoV-2-infection, primary analysis set. N1= Number of participants with non-missing data at baseline (injection 1/pre-booster) and the corresponding post-baseline timepoint.<br>†Number of participants with non-missing data at the timepoint (baseline or post-baseline).<br>§GMFR of nAb at day 29 post-baseline timepoint over pre-dose day 1 with corresponding 95% CI based on the t-distribution of log-transformed values or difference in the log-transformed values for GMT value and GMT fold-rise, respectively, then back transformed to the original scale.<br>¶ Log-transformed antibody levels are analyzed using an ANCOVA model with the treatment variable as fixed effect, adjusting for age group (<65, ≥65 years) and pre-booster titers. The resulting LS means, difference of LS means, and 95% CI are back transformed to the original scale.<br>\|\|95% CI was calculated by stratified Miettinen-Nurminen method adjusted by age group.<br>‡Seroresponse at a participant level defined as a change from <LLOQ to ≥4 × LLOQ, or at least a 4-fold rise if baseline is ≥LLOQ; comparison to pre-vaccination/pre-booster baselines. Percentages were based on the number of participants with non-missing data at baseline and the corresponding time point, 95% CI was calculated using the Clopper-Pearson method.<br>††The SRR difference is a calculated common risk difference using inverse-variance stratum weights and the middle point of Miettinen-Nurminen confidence limits of each one of the stratum risk differences. The stratified Miettinen-Nurminen estimate and the CI cannot be calculated when the seroresponse rate in both groups is 100%, absolute difference is reported.<br>¶¶ Exceeded non-inferiority criteria and met superiority criteria including lower bound CI >1 and testing sequence. |  |  |  |  |

To explore whether time intervals between the first and second booster doses influenced the post-booster neutralizing antibody titers, we analyzed the antibody responses across quartile time intervals for the mRNA-1273.222 and mRNA-1273 groups (participants with no previous infection, Table S7). Antibody titers did not appear to increase as the interval between prior doses increased, suggesting no evidence that these intervals had impact to the post-booster titers. Lastly, binding antibody levels across omicron BA.1, the ancestral SARS-CoV-2, alpha, beta, gamma, and delta variants were also higher following mRNA-1273.222 than mRNA-1273 boosters and GMRs (95% CI) ranged from 1.75 (1.57−1.96) to 2.03 (1.81−1.27) in participants with no prior infection (Table S8), and GMRs (95% CI) ranged from 1.52 (1.35−1.71) to 1.79 (1.58−2.03) in participants with prior infection (Table S9).

Given the emergence of the omicron BQ.1.1 and XBB.1 sublineages with the potential for immune escape, neutralization at day 29 against these variants was assessed in exploratory analyses for both omicron bivalent vaccines, mRNA-1273.222 and mRNA-1273.214 as described in the methods (Fig. S5 and Table S10). In mRNA-1273.222 participants without previous infection (n=40), pre-booster GMTs (95% CI) against BA.4/5, BQ.1.1 and XBB.1, respectively were 122.8 (74.3−203.1), 31.7 (19.6−51.3) and 18.1 (12.0−27.1); day 29 post-mbooster GMTs were 3355.4 (2109.9−5336.2), 621.9 (422.2−916.2) and 222.3 (147.4−335.2) with fold-rises (95% CI) from pre-booster titers of 27.3 (15.9−47.0), 19.6 (11.7−32.8), and 12.3 (7.4−20.5) for BA.4/5, BQ.1.1 and XBB.1. In mRNA-1273.222 participants with prior infection (n=20), pre-booster GMTs against BA.4/5, BQ.1.1 and XBB.1, respectively were 833.7 (422.5−1645.1), 124.7 (61.4−253.2) and 55.4 (28.4−108.0) and the day 29 post-booster GMTs were 8871.8 (4809.7−16364.8), 1093.5 (536.8−2227.9), and 381.4 (198.1−734.4), with fold-rises of 10.6 (6.4−17.6), 8.8 (5.0−15.5) and 6.9 (4.0−11.7) from pre-booster titers. Additionally, in the mRNA-1273.214 participants (n=40) with no evidence of previous infection, pre-booster GMTs against BA.4/5, BQ.1.1 and XBB.1, respectively were 121.1 (74.6−196.4), 39.2 (23.9−64.3) and 14.4 (9.5−21.7) and the day 29 post-booster GMTs were 602.1 (399.9−906.4), 161.1 (104.1−249.3) and 50.6 (32.4−79.2) with fold-rises of 5.0 (3.7−6.7), 4.1 (3.0−5.5) and 3.6 (2.5−5.1) from pre-booster titers. In the mRNA-1273.214 participants with prior infection (n=20), pre-booster GMTs against BA.4/5, BQ.1.1 and XBB.1, respectively were 1067.6 (506.2−2251.7), 147.8 (80.9−270.0) and 75.1 (35.4−159.1) and the post-booster GMTs were 3116.8 (1776.8−5467.3), 475.5 (304.7−742.0) and 214.2 (116.9−392.4) with fold-rises of 2.9 (1.9−4.4), 3.2 (2.3−4.5) and 2.9 (2.1−3.9) from pre-booster titers.

### Incidence of SARS-CoV-2 infections

In mRNA-1273.222 participants starting 14 days post-booster regardless of pre-booster SARS-CoV-2-infection status, 17 (3.3%) SARS-CoV-2 infections occurred with 9 (1.8%) asymptomatic infections and 8 (1.6%) Covid-19 events per the CDC definition (Table S11). There were no emergency room visits or hospitalizations due to these Covid-19 events. The incidence of SARS-CoV-2 infections and Covid-19 post-boost was comparable to the previously published incidence following the mRNA-1273 and mRNA-1273.214 booster doses.^5^

## DISCUSSION

In this study, the frequency of adverse reactions with the omicron BA.4/BA.5 containing bivalent booster mRNA-1273.222 was similar to or lower than that prior doses of mRNA-1273.^1,18^ The mRNA-1273.222 safety and reactogenicity data presented herein together with the previously published reactogenicity and 3-month follow-up safety information on the BA.1 bivalent booster^5,6^ mRNA-1273.214 add to our evaluation of the safety profile of omicron-targeting bivalent vaccines and both boosters have a safety profile similar to the original vaccine mRNA-1273.^1,18^ Additionally, the frequency of adverse reactions with mRNA-1273.222 does not appear to increase in participants with previous SARS-CoV-2 infection. Longer-term evaluation of the safety of mRNA-1273.222 continues in this ongoing study.

The 50-μg omicron BA.4/BA.5 bivalent vaccine mRNA-1273.222 elicited superior BA.4/BA.5 neutralizing antibody responses and non-inferior ancestral virus responses compared to 50-μg mRNA-1273 28 days after the booster dose, in participants without prior SARS-CoV-2 infection. Neutralizing antibody responses were also higher in mRNA-1273.222 participants with prior infection, compared to mRNA-1273, and no ceiling has been observed in the antibody titers in those with prior infection in this study and in previous evaluations.^5,22^ Additionally, the bivalent booster appears to similarly increase the antibody responses in participants 65 years-old, compared to younger participants, suggesting a consistent immunogenicity benefit in vulnerable populations. These results further support that an omicron-containing bivalent booster elicits potent responses against the variant contained in the vaccine with no decrement in the antibody response against the original virus.^5,6^ We have also previously reported that neutralizing antibody responses with an omicron BA.1 bivalent booster (mRNA-1273.214) are also more persistent compared to the original vaccine.^6^ Immunization with omicron-containing vaccines appear to elicit new germinal centers and de novo B-cell populations^7^ that are likely to contribute to improved neutralization potential. These benefits of the bivalent booster are likely to confer improved protection against Covid-19, relative to immunization with the original vaccine only, as emerging observation data suggest.^14,15^

Omicron variants continue to evolve with spike protein mutations, frequently clustered in the spike receptor binding and the N-terminal domains, that can confer immune escape. We assessed day 29 cross-neutralization against the emerging BQ.1.1 and XBB.1 omicron sublineages in mRNA-1273.222 recipients as well as in recipients of the BA.1 bivalent booster vaccine mRNA-1273.214.^5^ In recipients without prior infection, BQ.1.1 neutralizing antibody titers rose ∼20-fold and ∼4-fold and XBB.1 titers rose ∼12-fold and ∼4-fold, compared to pre-booster levels, for mRNA-1273.222 and mRNA-1273.214, respectively. Additionally, compared to the corresponding BA.4/BA.5 titers, BQ.1.1 titers were ∼4-5 fold lower and XBB.1 titers were ∼12-15 fold lower for both vaccines, consistent with the greater potential for immune escape by XBB.1. Neutralization was improved in participants with prior infection. These results underscore the cross-neutralization ability of omicron bivalent boosters against antigenically divergent variants.^5,6^

This study was not randomized and the immunogenicity for mRNA-1273.222 was compared to the within-study non-contemporaneous mRNA-1273 comparator given that it was no longer feasible to enroll mRNA-1273 participants after the authorization and approval of bivalent boosters. Because the study was conducted in response to the emerging epidemiology, we did not control for booster intervals. The interval between prior booster doses was longer in the mRNA-1273.222 (median 9.5 months) compared to mRNA-1273 (median 4.4 months) group. Although our analyses did not suggest an impact of interval on post-boost titers in either the mRNA-1273 or mRNA-1273.222, the intervals between the two groups were almost completely non-overlapping. This study was not designed to evaluate vaccine effectiveness; the incidence of SARS-CoV-2 infections within one month post-boost is nonetheless summarized and is similar to the previously reported incidence for bivalent and original vaccine groups;^5^ real-world vaccine effectiveness data is expected to further support the benefit of bivalent boosters.^14,15^ Lastly, the cross-neutralization assessment was performed with subgroups of study participants to enable rapid evaluation.

In conclusion, the omicron BA.4/BA.5-containing mRNA-1273.222 vaccine elicited superior BA.4/BA.5 neutralizing antibody responses compared to the original vaccine mRNA-1273 with no new safety concerns. These results are consistent with the omicron BA.1 bivalent vaccine mRNA-1273.214 which also induced potent and persistent neutralizing antibody responses as previously reported.^5,6^ Both omicron bivalent vaccines exhibited cross-neutralization against the emerging variants BQ.1.1 and XBB.1 despite their potential to evade immunity.^5^ Optimal vaccination strategies against Covid-19 will require continued variant surveillance and real-world vaccine effectiveness monitoring; it remains a priority to accelerate vaccine development at pace with viral evolution.

## Supporting information

Consort checklist

## Data Availability

As the trial is ongoing, access to patient-level data and supporting clinical documents by qualified external researchers may be available upon request and subject to review once the trial is complete.

## AUTHOR CONTRIBUTIONS

SC, JF, HZ, JMM and RD contributed to the design of the study and oversight. SC, JW, FE, BE, SK, PB, AB and NMcG contributed to data collection. BG, XS and DCM were responsible for immunogenicity assays. SC, JF, XZ, XC, HZ, RD, JMM, LRB and JET contributed to data analysis and/or interpretation of the data. SC, JF, and JET contributed to drafting the manuscript. All authors critically reviewed and provided input to manuscript drafts and approved the final version for submission to the journal.

## ACKNOWLEGEMENTS

We thank the participants in the trial and the members of the mRNA-1273 trial team (listed in the Supplementary Appendix) for their dedication and contributions to the trial, the Immune Assay Team at Duke University Medical Center, Durham, NC for PsVNA analyses, the SARS-CoV-2 Assessment of Viral Evolution (SAVE) program for help with variant reagent development and Frank J. Dutko, Ph.D., (Moderna consultant) for figure development and editorial support.

