## Supplementary Material for "Safety and Immunogenicity of Omicron BA.4/BA.5 Bivalent Vaccine Against Covid-19"

**Supplementary Appendix**

**TABLE OF CONTENTS**


| **Contents** | **Pg.** |
|---|---|
| List of Investigators | 3 |
| Supplementary Methods | 4 |
| Study Design | 4 |
| Trial Registration | 4 |
| Trial Oversight | 4 |
| Part H Study Eligibility Criteria | 5 |
| Trial Vaccine | 6 |
| Immunogenicity Objectives and Endpoints | 7 |
| Statistical Analysis of Immunogenicity | 7 |
| Incidence of SARS-CoV-2 Infection and Covid-19 | 10 |
| Immunogenicity assays | 11 |
| Figure S1. Trial Profile of Analysis Sets | 14 |
| Figure S2: Statistical Testing Sequence for Immunogenicity Primary Objectives | 15 |
| Figure S3. Distribution of Observed Neutralizing Antibody Titers Against Omicron BA.4/BA.5 and SARS-CoV-2 (D614G) after 50-µg of mRNA-1273.222 and mRNA-1273 Administered as Second Booster Doses in Participants With and Without Prior SARS-CoV-2 Infection | 16 |
| Figure S4. Observed Neutralizing Antibody Response Omicron BA.4/BA.5 and Ancestral SARS-CoV-2 (D614G) after 50-µg of mRNA-1273.222 and mRNA-1273 Administered as Second Boosters in Participants with No Prior SARS-CoV-2-Infection by Age Groups | 18 |
| Figure S5. Observed Neutralizing Antibody Titers Against Omicron Variants after 50-µg of mRNA-1273.222 and mRNA-1273.214 Administered as Second Booster Doses by Prior SARS-CoV-2 Infection Status at Pre-Booster | 19 |
| Table S1. Objectives and Endpoints for Part H Second Booster Doses of 50-µg mRNA-1273.222 in Participants Who Received 100 µg mRNA-1273 Primary Series and a Booster Dose of 50 µg mRNA-1273 | 21 |
| Table S2. Analysis Sets | 23 |
| Table S3. Demographics for Subset of Participants Included in BQ1.1 and XBB.1 Variant Analysis. | 24 |
| Table S4. Solicited Local and Systemic Adverse Reactions Within 7 Days Following the Second Booster Injections of mRNA-1273.222 (50 µg) by Pre-booster SARS-CoV-2 Status, Solicited Safety Set | 25 |
| Table S5. Summary of Unsolicited Adverse Events ≤28 Days Post-booster Dose and Up to Data Cutoff, Safety Set | 27 |
| Table S6. Geometric Mean Neutralizing Antibody Titer Ratio and Seroresponse Difference Against Ancestral SARS-CoV-2 (D614G) and Omicron BA.4/BA.5 after 50-µg of mRNA-1273.222 and mRNA-1273 administered as Second Booster Doses in Participants with Evidence of SARS-CoV-2 Infection at Pre-booster | 28 |
| Table S7. Neutralizing Omicron BA.4/BA.5 Neutralizing Titers by Time Intervals Between First Booster Dose of mRNA-1273 and Second Booster Doses of mRNA-1273.222 and mRNA-1273 in Participants Without Prior Infection | 29 |
| Table S8. Observed Binding Antibody Titers and Geometric Mean Ratios Against Ancestral SARS-CoV-2 (D614G) and Variants after 50-µg of mRNA-1273.222 and | 30 |

| | |
|---|---|
| mRNA-1273 Administered as Second Booster Doses in Participants with No Evidence of SARS-CoV-2 Infection at Pre-Booster | |
| Table S9. Observed Binding Antibody Levels and Geometric Mean Ratios Against Ancestral SARS-CoV-2 and Variants After Second 50-μg Boosters of mRNA-1273.222 and mRNA-1273 in Participants with Evidence of SARS-CoV-2 Infection at Pre-booster | 31 |
| Table S10. Observed Neutralizing Antibody Geometric Mean Titers Against Omicron Variants BA.4/BA.5, BQ1.1 and XBB.1 after 50-μg of mRNA-1273.222 and mRNA-1273.214 Administered as Second Booster Doses by Prior SARS-CoV-2 Infection Status at Pre-Booster | 32 |
| Table S11. Incidences of SARS-CoV-2-infection and Covid-19 Starting 14 Days Post-booster | 33 |
| Supplementary References | 34 |

### List of Investigators

| Investigator | Institution | Location |
|---|---|---|
| Paul Bradley | Meridian Clinical Research | Savannah, Georgia |
| Adam Brosz | Meridian Clinical Research | Grand Island, Nebraska |
| Laurence Chu | Benchmark Research | Austin, Texas |
| Michael Cotugno | Benchmark Research | Metairie, Louisiana |
| Frank Eder | Meridian Clinical Research, LLC | Binghamton, New York |
| Brandon Essink | Meridian Clinical Research | Omaha, Nebraska |
| Carlos Fierro | Johnson County Clin-Trials | Lenexa, Kansas |
| Veronica Fragoso | DM Clinical Research - Texas Center for Drug Development | Houston, Texas |
| Carl Griffin | Lynn Health Science Institute | Oklahoma City, Oklahoma |
| Greg Hachigian | Benchmark Research | Sacramento, California |
| Shishir Khetan | Meridian Clinical Research | Rockville, Maryland |
| Michael Koren | Jacksonville Center for Clinical Research | Jacksonville, Florida |
| Vicki Miller | DM Clinical Research | Tomball, Texas |
| Paul Pickrell | Tekton Research Inc. | Austin, Texas |
| Rachel Presti | Infectious Disease Clinical Research | Saint Louis, Missouri |
| Howard Schwartz | Research Centers of America | Hollywood, Florida |
| William Seger | Benchmark Research | Fort Worth, Texas |
| Charles Harper. Keith Vrbicky, | Meridian Clinical Research | Norfolk, Nebraska |
| Larkin Wadsworth | Sundance Clinical Research | Saint Louis, Missouri |
| Stephen Walsh | Brigham and Womens Hospital | Boston, Massachusetts |
| Jordan Whatley | Meridian Clinical Research | Baton Rouge, Louisiana |

### SUPPLEMENTARY METHODS

#### Study Design

This is an open-label, ongoing, phase 2/3 study to evaluate the immunogenicity, safety, and reactogenicity of various modified mRNA-1273 vaccine candidates against Covid-19 administered as boosters (mRNA-1273.211 [Part A.1], mRNA-1273 [Part B], mRNA-1273.617.2 [Part C], mRNA-1273.213 [Part D and E], mRNA-1273.529 [Part F], mRNA-123.214 [Part G and Part A.2], and mRNA-1273.222 [Part H] vaccines; clintrials.gov NCT04927065). Part H interim results are reported here. In part H, the bivalent mRNA-1273.222 vaccine which contains 25 µg each of two mRNAs encoding the ancestral SARS-CoV-2 and omicron variant (BA.4/BA.5) spike sequences is tested as a second booster dose at 50-µg in comparison to mRNA-1273 given as a second booster dose (part F, cohort 2, 50-µg mRNA-1273).

Enrollment of the mRNA-1273.222 50-µg second boost arm was initiated August 10, 2022. Part H evaluates the immunogenicity, safety, and reactogenicity of 50-µg of the mRNA-1273.222 vaccine candidate when administered as a second booster dose to adults who have previously received 2 doses of 100 µg mRNA-1273 as a primary series and a first booster dose of mRNA-1273 (50 µg) in the Coronavirus Efficacy (COVE) trial[1,2] or under US emergency use authorization (EUA). The immunogenicity of mRNA-1273.222 50-µg is compared to that induced after the second booster dose of mRNA-1273 (Part F, cohort 2, 50-µg mRNA-1273). Participants were enrolled in the within-study non-contemporaneous mRNA-1273 group (Part F, cohort 2) during February 18th-March 8th, 2022 and data for this group is based on the data cutoff date of July 6, 2022 at the day 91 interim analysis.[3] Participants were enrolled during August 10-23 for mRNA-1273.222 (Part H).

#### Trial Registration

The study protocol was approved by the Institutional Review Board on May 26th 2021, and enrollment in part A of the study was initiated on May 28, 2021, before the trial posting date of June 14, 2021 on the clintrials.gov registration website as we were emergently advancing Covid-19 booster vaccine candidates to combat the SARS-CoV-2 pandemic (the objective of this trial). Nonetheless, the trial was registered within 21 days after the first participant was enrolled, compliant per clintrials.gov guidance (https://clinicaltrials.gov/ct2/manage-recs/fdaaa). Additionally, in parts F and H of the study presented in this manuscript, the 890 participants were enrolled during August 10th-23rd, 2022 for mRNA-1273.222 (Part H) and during February 18th-March 8th, 2022 (Part F, cohort 2) for mRNA-1273. Moderna has received an emergency use authorization (EUA) of our booster vaccine mRNA-1273 by regulatory agencies, and we are also currently pursuing EUA for booster candidates evaluated in this trial.

#### Trial Oversight

The trial is being conducted across 23 US sites, in accordance with the International Council for Harmonisation of Technical Requirements for Registration of Pharmaceuticals for Human Use, Good Clinical Practice guidelines. The central Institutional Review Board approved the protocol and consent forms. All participants provided written informed consent.

SC, JF, JMM, RD and HZ contributed to the design of the study and oversight. SC, JW, FE, BE, SK, PB, AB and NMcG contributed to data collection. BG and DCM were responsible for immunogenicity assays. SC, JF, XZ, XC, HZ, RD, JMM, LRB and JET contributed to data analysis and/or interpretation of the data. SC, JF, and JET contributed to drafting the manuscript.

All authors critically reviewed and provided input to manuscript drafts and approved the final version for submission to the journal.

**Part H Study Eligibility Criteria**
***Inclusion Criteria:***

Each participant must meet all of the following criteria to be enrolled in this study:

1. Male or female, at least 18 years of age at the time of consent (Screening Visit).
2. Investigator's assessment that participant understands and is willing and physically able to comply with protocol-mandated follow-up, including all procedures.
3. Participant has provided written informed consent for participation in this study, including all evaluations and procedures as specified in this protocol.
4. Female participants of nonchildbearing potential may be enrolled in the study. Nonchildbearing potential is defined as surgically sterile (history of bilateral tubal ligation, bilateral oophorectomy, hysterectomy) or postmenopausal (defined as amenorrhea for ≥ 12 consecutive months prior to Screening [day 0] without an alternative medical cause). A follicle-stimulating hormone level may be measured at the discretion of the investigator to confirm postmenopausal status.
5. Female participants of childbearing potential may be enrolled in the study if the participant fulfills all of the following criteria:
   - Has a negative pregnancy test on the day of vaccination (day 1)
   - Has practiced adequate contraception or has abstained from all activities that could result in pregnancy for at least 28 days prior to day 1
   - Has agreed to continue adequate contraception through 3 months following vaccination.
   - Is not currently breastfeeding

   (Adequate female contraception is defined as consistent and correct use of a Food and Drug Administration approved contraceptive method in accordance with the product label)
6. Participant must have been either previously enrolled in the phase 3 mRNA-1273 COVE trial,[1,2] must have received 2 doses of mRNA 1273 in that study, with his/her second dose at least 6 months prior to enrollment in this study, and must be currently enrolled and compliant in that study (i.e., has not withdrawn or discontinued early); or participant must have received 2 doses of mRNA-1273 under the EUA with their second dose at least 6 months prior to enrollment in this study; or have received a 2 dose primary series of mRNA-1273 followed by a 50-µg booster dose of mRNA-1273 in the mRNA-1273 COVE trial or under EUA at least 3 months prior to enrollment this study; and able to provide proof of vaccination status at the time of screening (day 1).

***Exclusion Criteria:***

Participants meeting any of the following criteria at the Screening Visit, unless noted otherwise, will be excluded from the study:

1. Had significant exposure to someone with SARS-CoV-2 infection or coronavirus disease 2019 (Covid-19) in the past 14 days, as defined by the CDC as a close contact of someone who has Covid-19).
2. Has known history of SARS-CoV-2 infection within 3 months prior to enrollment.

3. Is acutely ill or febrile (temperature ≥38.0°C [100.4°F]) less than 72 hours prior to or at the Screening Visit or day 1. Participants meeting this criterion may be rescheduled and will retain their initially assigned participant number.
4. Currently has symptomatic acute or unstable chronic disease requiring medical or surgical care, to include significant change in therapy or hospitalization for worsening disease, at the discretion of the investigator.
5. Has a medical, psychiatric, or occupational condition that may pose additional risk as a result of participation, or that could interfere with safety assessments or interpretation of results according to the investigator's judgment.
6. Has a current or previous diagnosis of immunocompromising condition to include human immunodeficiency virus, immune-mediated disease requiring immunosuppressive treatment, or other immunosuppressive condition.
7. Has received systemic immunosuppressants or immune-modifying drugs for >14 days in total within 6 months prior to Screening (for corticosteroids ≥10 mg/day of prednisone equivalent) or is anticipating the need for immunosuppressive treatment at any time during participation in the study.
8. Has known or suspected allergy or history of anaphylaxis, urticaria, or other significant AR to the vaccine or its excipients.
9. Has a documented history of myocarditis or pericarditis within 2 months prior to Screening Visit (day 0).
10. Coagulopathy or bleeding disorder considered a contraindication to intramuscular (IM) injection or phlebotomy.
11. Has received or plans to receive any licensed vaccine ≤ 28 days prior to the injection (day 1) or a licensed vaccine within 28 days before or after the study injection, with the exception of influenza vaccines, which may be given 14 days before or after receipt of a study vaccine.
12. Has received systemic immunoglobulins or blood products within 3 months prior to the Screening Visit (day 0) or plans for receipt during the study.
13. Has donated ≥450 mL of blood products within 28 days prior to the Screening Visit or plans to donate blood products during the study.
14. Plans to participate in an interventional clinical trial of an investigational vaccine or drug while participating in this study.
15. Is an immediate family member or household member of study personnel, study site staff, or Sponsor personnel.
16. Is currently experiencing an SAE in COVE trial at the time of screening for this study.

**Trial Vaccine**

The mRNA-1273.222 50-µg vaccine contains equal amounts of mRNAs (25-µg of each mRNA sequence) that encode the prefusion-stabilized spike glycoproteins of the ancestral SARS-CoV-2 (Wuhan-Hu-1) and the omicron variants (BA.4 and BA.5, and having identical sequences are deignated as BA.4/BA.5). The mRNA-1273 50-µg vaccine (Moderna Covid-19 Vaccine) contains only the mRNA sequence encoding the spike glycoprotein of the ancestral SARS-CoV-2. In both vaccines, mRNAs are encapsulated in lipid nanoparticles (LNPs) as described previously.[4] The booster doses of mRNA-1273.222 and mRNA-1273 were administered by intramuscular injection at doses of 50-µg of mRNA in a 0.5 mL volume.

**Immunogenicity Objectives and Endpoints (Part H)**
There were four pre-specified primary immunogenicity objectives at day 29 for part H in the study (Table S1 and Fig. S2).

- To demonstrate non-inferiority of the antibody response of a second booster dose of mRNA-1273.222 50 µg compared to mRNA-1273 50 µg when administered as a second booster dose against the omicron BA.4/5 based on GMT ratio and SRR difference at day 29.
- To demonstrate superiority of the antibody response of a second booster dose of mRNA-1273.222 compared to mRNA-1273 (50 µg) administered as a second booster dose against the omicron BA.4/5 based on GMT ratio at day 29.
- To demonstrate non-inferiority of the antibody response of mRNA-1273.222 50 µg compared to mRNA-1273 50 µg when administered as a second booster dose against the ancestral SARS-CoV-2 D614G based on GMT ratio and SRR difference at day 29.

Endpoints for the primary objectives were:

- GMT ratio of omicron BA.4/5 GMT of mRNA-1273.222 over the omicron BA.4/5 GMT of mRNA-1273 (Part F, Cohort 2, 50 µg mRNA-1273) at Day 29
- SRR difference between mRNA-1273.222 against omicron BA.4/5 and mRNA-1273 against omicron BA.4/5 at day 29
- GMT ratio of the ancestral SARS-CoV-2 D614G GMT of mRNA-1273.222 over the ancestral SARS-CoV-2 D614G GMT of mRNA-1273 (Part F, Cohort 2, 50 µg mRNA-1273) at day 29
- SRR difference between mRNA-1273.222 and mRNA-1273 against the ancestral SARS-CoV-2 D614G at day 29

A secondary objective is to evaluate the immunogenicity of mRNA-1273.222 50 µg as a second booster dose against the ancestral SARS-CoV-2 (and other variants) compared to a second booster dose of mRNA-1273 50 µg at all timepoints post-booster. The secondary endpoints were GMT ratio of mRNA-1273.222 50 µg and mRNA-1273 50 µg against ancestral SARS-CoV-2 (and other variants) at all timepoints post-booster and the SRR difference between mRNA-1273.222 50 µg and mRNA-1273 50 µg against ancestral SARS-CoV-2 (and other variants) at all timepoints post-booster.

**Statistical Analysis of Immunogenicity**
In part H of the study, 50-µg mRNA-1273.222 as the second booster dose is compared to 50-µg mRNA-1273 as the second booster dose (active control arm in part F, Cohort 2).[3] For the primary objective on immune response, 5 hypotheses are specified to be evaluated at day 29 post-booster (online protocol and SAP and Fig. S2). Interim analysis results for the day 29 hypotheses are presented in this report. Below are the 5 hypotheses for day 29:

a. H1: 50 µg mRNA 1273.222, as a second booster dose, against omicron BA.4/5 is noninferior to the second booster dose of (50 µg) mRNA 1273 against omicron BA.4/5 based on the GMT ratio of mRNA 1273.222 against omicron BA.4/5 at Day 29 compared to mRNA 1273 against omicron BA.4/5 at Day 29 with a non-inferiority margin of 1.5.
b. H2: 50 µg mRNA-1273.222, as a second booster dose, against omicron BA.4/5 is non-inferior to the second booster dose of (50 µg) mRNA-1273 against omicron BA.4/5 based on the difference in SRR at Day 29 with a non-inferiority margin, if
    - o the lower bound of the 95% CI of the SRR difference (50 µg mRNA-1273.222 against omicron BA.4/5 at Day 29 - 50 µg mRNA-1273 against omicron BA.4/5 at Day 29) is >-

5%, then the non-inferiority of mRNA-1273.222 against omicron BA. 4/5 compared to that of mRNA-1273 is demonstrated based on a non-inferiority margin of 5%.

  - the lower bound of the 95% CI of the SRR difference (50 µg mRNA-1273.222 against omicron BA.4/5 at day 29 - 50 µg mRNA-1273 against omicron BA.4/5 at day 29) is >-10% but ≤ -5%, then non-inferiority of mRNA-1273.222 against omicron BA. 4/5 compared to that of mRNA-1273 is demonstrated based on a non-inferiority margin of 10%.

c. H3: 50 µg mRNA-1273.222, as a second booster dose, against the ancestral SARS-CoV-2 D614G is non-inferior to the second booster dose of (50 µg) mRNA-1273 against the ancestral SARS-CoV-2 D614G based on the GMT ratio of mRNA-1273.222 against the ancestral SARS-CoV-2 D614G at day 29 compared to mRNA-1273 against the ancestral SARS-CoV-2 D614G at day 29 with a non-inferiority margin of 1.5.
d. H4: 50 µg mRNA-1273.222, as a second booster dose, against the ancestral SARS-CoV-2 D614G is non-inferior to the second booster dose of (50 µg) mRNA-1273 against ancestral SARS-CoV-2 D614G based on the difference in SRR at day 29 with a non-inferiority margin 10%.
e. H5: 50 µg mRNA-1273.222, as a second booster dose, against the omicron BA.4/BA.5 is superior to the second booster dose of (50 µg) mRNA-1273 against omicron BA.4/BA.5 day 29.

For the primary immunogenicity objective, an alpha of 0.05 (two-sided) is allocated to the day 29 time point for hypotheses testing. The primary immunogenicity objective is considered met for non-inferiority when the lower bound of the 95.0% confidence interval (CI) of GMR is >0.667 and the seroresponse rate-difference is >-10%. Superiority is considered met when the lower bound of the 95.0% CI of GMR is >1. These non-inferiority and superiority criteria were chosen based on FDA guidelines.[5,6] Day 29 interim analysis results are presented. The within-study non-contemporaneous mRNA-1273 comparator group (Part F, cohort 2) was enrolled during February 18th-March 8th, 2022 and data for this group, based on the data cutoff date of July 6, 2022 at the day 91 interim analysis, is used for the immunogenicity comparison.

***Sample Size***

The target enrollment is approximately 500 participants for 50 µg mRNA 1273.222. Assuming 40% of participants are excluded from the PP Set for Immunogenicity – SARS-CoV-2 negative (due to a SARS-CoV-2 infection pre-booster), with approximately 300 participants in 50 µg mRNA-1273.222 and 260 participants in 50 µg mRNA-1273 (Part F, Cohort 2-50 µg mRNA-1273) in the PP Set for Immunogenicity and SARS-CoV-2 negative, there is approximately 60% power to demonstrate the primary immunogenicity objectives with an alpha of 0.05 (2-sided) at day 29. The assumptions are: the true GMR (mRNA-1273.222 second booster vs. mRNA-1273 second booster) against omicron BA.4/5 is 1.5, GMR (mRNA-1273.222 second booster vs. mRNA-1273 second booster) against ancestral SARS-CoV-2 D614G is 1, the standard deviation of the log-transformed titer is 1.5, and the non-inferiority margin for GMR is 1.5. The true SRR against omicron BA.4/5 after mRNA-1273.222 as a second booster dose is 95% (same assumption for 50 µg mRNA-1273), and non-inferiority margin for SRR difference against omicron BA.4/5 is 5%. The true SRR against ancestral SARS-CoV-2 D614G after mRNA-1273.222 as a second booster dose is 95% (same assumption for 50 µg mRNA-1273), and the non-inferiority margin for SRR difference against ancestral SARS-CoV-2 D614G is 10%.

An analysis of covariance (ANCOVA) model was performed to assess the difference in antibody responses between mRNA-1273.222 and mRNA-1273 booster doses, with antibody titers post-booster as a dependent variable, and a group variable (mRNA-1273.222 and mRNA-1273) as the fixed effect, adjusting for age groups (<65, ≥65 years) and pre-booster antibody titers. The GMTs (95% CI) estimated by the geometric least square mean (GLSM) from the model for each group and the GMR (mRNA-1273.222 compared with mRNA-1273) estimated by the ratio of GLSM from the model (95.0% CIs) are provided. The 95.0% CI for GMR was used to assess the between group difference in antibody responses.

Seroresponse is defined as ≥4 × lower limit of quantification (LLOQ) for those with baseline < LLOQ; ≥4-fold-rise for those baseline ≥LLOQ. Seroresponse is derived based on two types of baselines:

1) Pre-vaccination (pre-injection 1 of the primary series)

2) Pre-booster baseline

Both definitions were used when comparing seroresponse. Seroresponse, based on change (fold-rise) from pre-injection 1 of the primary series, was considered the primary approach for seroresponse.

For participants without pre-injection 1 antibody titer information, seroresponse is defined as ≥4X LLOQ for those with negative SARS-CoV-2 status at pre-injection 1 of the primary series, and antibody titers are imputed as <LLOQ at pre-injection 1 of primary series. For participants who are without SARS-CoV-2 status information at pre-injection 1 of primary series, their pre-booster SARS-CoV-2 status is used to impute their SARS-CoV-2 status at their pre-injection 1 of primary series.

The SRR of each arm against ancestral SARS-CoV-2 (D614G) and variants, defined as the percentage of participants achieving SRR against ancestral SARS-CoV-2 (D614G) and variants respectively, are provided for each arm with the 95% CI calculated using the Clopper Pearson method. The differences of SRR between mRNA-1273.222 and mRNA-1273 are calculated with 95.0% CI based on stratified Miettinen-Nurminen method adjusting for age groups.

An analysis of the primary immunogenicity endpoints was also performed in the per-protocol set for immunogenicity (participants with and without evidence of prior SARS-CoV-2 infection pre-booster), using an ANCOVA model, with antibody titers at day 29 post-booster as the dependent variable and the vaccine group variable as the fixed effect, adjusting for age groups (<65, ≥65 years), pre-booster SARS-CoV-2 infection status, and pre-booster titers. The SRR difference between the mRNA-1273.2224 and mRNA-1273 groups was calculated with 95.0% CI based on stratified Miettinen-Nurminen method adjusted for the pre-booster SARS-CoV-2 infection status and age group. A pre-planned subgroup analysis of participants with prior evidence of SARS-CoV-2-infection pre-booster was performed using an ANCOVA model to assess neutralizing antibody differences between the mRNA-1273.222 and mRNA-1273 groups based on GMRs with 95% CIs. Lastly, a sensitivity analysis was performed excluding the participants with evidence of SARS-CoV-2-infection after the booster dose.

Observed binding antibody GMTs and 95% CIs against variants are provided. Binding antibody level differences between the mRNA-1273.222 and mRNA-1273 groups based on GMRs with 95% CIs were assessed using an ANCOVA model, adjusting for age group and pre-booster titers.

Immunogenicity against emerging omicron BQ.1.1 and XBB.1 variants was additionally explored in random samples of recipients (n=60) stratified by age and baseline pre-booster titers from the full analysis sets of the mRNA-1273.222 group and from the previously described omicron BA.1-containing bivalent booster mRNA-1273.214 group at NEJM.org (Table S3).[5] Among those with negative SARS-CoV-2 pre-booster status, participants were first stratified by age (every 5 years from 20-90 years) and by deciles of baseline pre-booster BA.4/BA.5 titers, then randomly selected as 40 pairs (n=40/each group; n=20 each from age groups <65 years and ≥65) from two booster groups. Participants SARS-CoV-2 positive were stratified by age (every 5 years from 20-90 years) and by quartiles of baseline pre-booster BA.4/BA.5 titers, then randomly selected in 20 pairs in each booster group (n=20; n=10 each from age groups <65 years ≥65).

| **Sampling method** | **SARS-COV-2 Negative n=40** | | | **SARS-COV-2 Positive n=20** | | |
|---|---|---|---|---|---|---|
| **Baseline Covariate Strata*** | **age (every 5 years from 20-90 years) and baseline pre-booster BA.4/BA.5 titer deciles** | | | **age (every 5 years from 20-90 years) and baseline pre-booster BA.4/BA.5 titer quartiles** | | |
| | **18-64 yr** | **≥65 yr** | **All** | **18-64 yr** | **≥65 yr** | **All** |
| **mRNA-1273.222 (n)** | 20 | 20 | 40 | 10 | 10 | 20 |
| **mRNA-1273.214 (n)** | 20 | 20 | 40 | 10 | 10 | 20 |

Pseudovirus neutralizing antibodies against BQ.1.1 and XBB.1 were assessed in subsets of recipients (n=60) stratified by age and baseline pre-booster titers from the mRNA-1273.222 group and from the previously described omicron BA.1-containing bivalent booster mRNA-1273.214 group at NEJM.org (Table S3) in the full analysis sets.[5] *Among those with negative SARS-CoV-2 pre-booster status, participants were first stratified by age (every 5 years from 20-90 years) and by deciles of baseline pre-booster BA.4/BA.5 titers and participants SARS-CoV-2 positive were stratified by age (every 5 years from 20-90 years) quartiles of baseline pre-booster BA.4/BA.5 titers. Pairs of participants were then randomly selected from age groups <65 years ≥65 for the analysis.

### Incidence of SARS-CoV-2 Infection and Covid-19

Vaccine effectiveness was not assessed in this trial, but Covid-19 and SARS-CoV-2 infection were actively surveilled through weekly contact and blood draws. An exploratory objective of the study is to assess symptomatic and asymptomatic SARS-CoV-2 infection. SARS-CoV-2 infection is a combination of symptomatic infection (Covid-19) and asymptomatic SARS-CoV-2 infection for participants with negative SARS-CoV-2 status pre-booster. Symptomatic infection was evaluated using the primary case definition in the COVE study[1,2] as well as a secondary case definition based on the Centers for Disease Control and Prevention (CDC) criteria.[7] Asymptomatic SARS-CoV-2 infection was defined as a positive reverse-transcriptase polymerase chain reaction (RT-PCR) test or a positive serologic test for anti-nucleocapsid antibody after a negative test at the time of enrollment, in the absence of symptoms.

#### *SARS-CoV-2 infection*

SARS-CoV-2 infection is defined in participants with negative SARS-CoV-2 status pre-booster by either binding antibody (bAb) levels against SARS-CoV-2 nucleocapsid protein negative (as measured by Roche Elecsys) at day 1 that becomes positive (as measured by Roche Elecsys) starting at day 29 or later, OR a positive RT-PCR counted starting 14 days after the booster dose.

For the analysis, documented infection is counted starting 14 days after the booster dose, which requires positive serology test result based on bAb specific to SARS-CoV-2 nucleocapsid at day 29 or later, or a positive RT-PCR result starting 14 days after the booster dose. The date of documented infection is the earlier of the date of a positive post-baseline RT-PCR result or of positive serology test result based on bAb specific to SARS-CoV-2 nucleocapsid. The time to first SARS-CoV-2 infection is calculated as the date of the 1st documented infection minus the date of injection + 1. Cases are counted starting 14 days after the injection (date of documented infection minus date of the injection ≥14). SARS-CoV-2 infection cases are summarized based on tests performed at least 14 days after the booster dose.

***Asymptomatic SARS-CoV-2 Infection***

Asymptomatic SARS-CoV-2 infection is measured by RT-PCR of nasal swabs and/or serology tests obtained at post-baseline study visits counted starting 14 days after the injection in participants with negative SARS-CoV-2 status pre-booster. Asymptomatic SARS-CoV-2 infection is identified by the absence of symptoms and infections as detected by RT-PCR or serology tests. Specifically, the absence of Covid-19 symptoms AND at least either a positive serology test result based on bAb specific to SARS-CoV-2 nucleocapsid protein day 29 or later, when blood samples for immunogenicity are collected, or a positive RT-PCR test at scheduled or unscheduled/illness visits. The date of documented asymptomatic infection is the earlier date of positive serology test result based on bAb specific to SARS-CoV-2 nucleocapsid due to infection, or positive RT-PCR, with absence of symptoms. The time to the asymptomatic SARS-CoV-2 infection is calculated as the date of asymptomatic SARS-CoV-2 infection minus the date of injection + 1.

***Symptomatic SARS-CoV-2 Infection (Covid-19)***

Symptomatic SARS-CoV-2 Infection (Covid-19) is defined as the incidence of the first occurrence of symptomatic SARS-CoV-2 infection measured by RT-PCR of nasal swabs starting 14 days after the booster dose. Surveillance for Covid-19 symptoms is conducted via weekly contact and blood draw, and an illness visit to collect a nasopharyngeal swab was arranged for participants reporting Covid-19 symptoms.

Two definitions of symptomatic SARS-CoV-2 infection (Covid-19). This includes the primary case definition in the COVE trial[1,2] based on a positive post-baseline RT-PCR result AND at least TWO systemic symptoms (fever (≥ 38ºC/≥ 100.4ºF), chills, muscle and/or body aches [not related to exercise], headache, sore throat, new loss of taste/smell; OR at least ONE of respiratory signs/symptoms (cough, shortness of breath and/or difficulty breathing, OR clinical or radiographical evidence of pneumonia). The second case definition is based on CDC criteria for symptomatic disease defined as a positive post-baseline RT-PCR test AND at least ONE systemic or respiratory symptoms (fever [≥ 38ºC/≥100.4ºF], chills, cough, shortness of breath and/or difficulty breathing, fatigue, muscle and/or body aches [not related to exercise], headache, new loss of taste/smell, sore throat, congestion, runny nose, nausea, vomiting, or diarrhea).[7]

The date of a documented Covid-19 case is the later date of a symptom and the date of positive RT-PCR test, and the two dates should be within 14 days of each other. The time to the first occurrence of Covid-19 is calculated as the date of documented Covid-19 minus the date of injection + 1. Cases are counted starting 14 days after the injection (date of documented Covid-19 minus date of the injection ≥14).

### Immunogenicity Assays

#### *SARS-CoV-2 Spike-Pseudotyped Virus Neutralization Assay*

SARS-CoV-2 neutralizing antibodies (nAb) in samples were assessed using a SARS-CoV-2 Spike (S)-Pseudotyped Virus Neutralization Assay (PsVNA) in 293/ACE2 cells.[8] The PsVNA quantifies nAb using lentivirus particles that express full-length spike proteins on their surface, and contain a firefly luciferase reporter gene for quantitative measurements of infection in transduced 293T cells expressing high levels of ACE2 (293T/ACE2 cells) by relative luminescence units (RLU). Serial dilution of antibodies was used to produce a dose-response curve. Neutralization was measured as the serum dilution at which RLU was reduced by 50% (ID50) relative to mean RLU in virus control wells (cells + virus but no sample) after subtraction of mean RLU in cell control wells (cells only). Positive controls were included on each assay plate in order to follow stability over time. The assay is validated for the prototypic D614G variant and the omicron BA.5/BA.5 sublineage but has not been validated for the omicron BQ.1.1 and XBB.1 sublineages.

The spike-pseudotyped viruses were derived from ancestral Wuhan-Hu-1 with the following amino acid substitutions: (prototype [D614G]; omicron subvariants BA.4 and BA.5 (designated BA.4/BA.5 for identical spike sequences between BA.4 and BA.5 [T191, L24S, ΔP25, ΔP26, ΔA27, G142D, V213G, G339D, S371F, S373P, S375F, T376A, D405N, R408S, K417N, N440K, L452R, S477N, T478K, E484A, F486V, Q498R, N501Y, Y505H, D614G, H655Y, N679K, P681G, N764K, D796Y, Q954H, N969K]); omicron subvariant BQ.1.1 [T19I, L24S, ΔP25, ΔP26, ΔA27, H69-, V70-, G142D, V213G, G339D, R346T, S371F, S373P, S375F, T376A, D405N, R408S, K417N, N440K, K444T, L452R, N460K, S477N, T478K, E484A, F486V, Q498R, N501Y, Y505H, D614G, H655Y, N679K, P681H, N764K, D796Y, Q954H, N969K]; omicron subvariant XBB.1 [T19I, L24S, ΔP25, ΔP26, ΔA27, V83A, G142D, Y145Q, ΔH146, Q183E, V213E, G252V, G339H, R346T, L368I, S371F, S373P, S375F, T376A, D405N, R408S, K417N, N440K, V445P, G446S, N460K, S477N, T478K, E484A, F486S, F490S, Q498R, N501Y, Y505H, D614G, H655Y, N679K, P681H, N764K, D796Y, Q954H, N969K].

#### *SARS-CoV-2 Meso Scale Discovery (MSD) assay*

The validated Meso Scale Discovery (MSD, Rockville, MD) assay (SARSCOV2S2P [VAC123]; https://www.mesoscale.com/products/v-plex-sars-cov-2-panel-24-igg-kit-k15575u/) uses an indirect, quantitative, electrochemiluminescence method to detect SARS-CoV-2 binding IgG antibodies that bind to the SARS-CoV-2 full-length spike protein (Wuhan-Hu-1 ancestral SARS-CoV-2; beta [B.1.351] with the following amino acid changes in the spike protein [L18F, D80A, D215G, Δ242-244, R246I, K417N, E484K, N501Y, D614G, and A701V]; alpha [B.1.1.7] with the following amino acid changes in the spike protein [ΔH69-V70, ΔY144, N501Y, A570D, D614G, P681H, T761I, S982A, and D1118H]; gamma [P.1] with the following amino acid changes in the spike protein [L18F, T20N, P26S, D138Y, R190S, K417T, E484K, N501Y, D614G, H655Y, T1027I, and V1176F]); delta [B.1.617.2; AY.4; Alt Seq 2] with the following amino acid changes in the spike protein [T19R, T95I, G142D, Δ156/157, R158G, L452R, T478K, D614G, P681R, and D950N]); omicron [B.1.1.529; BA.1] with the following amino acid changes in the spike protein [A67V, ΔH69-V70, T95I, G142D, Δ143-145, Δ211/L212I, ins214EPE, G339D, S371L, S373P, S375F, K417N, N440K, G446S, S477N, T478K, E484A, Q493R, G496S, Q498R, N501Y, Y505H, T547K, D614G, H655Y, N679K, P681H, N764K, D796Y, N856K, Q954H, N969K, and L981F]) in human serum. The assay was performed by

PPD, (Thermo Fisher Scientific Vaccines Laboratory Services, Richmond, Virginia). The assay is based on MSD technology which employs capture molecule MULTI-SPOT® microtiter plates fitted with a series of electrodes.

### Figure S1. Trial Profile of Analysis Sets

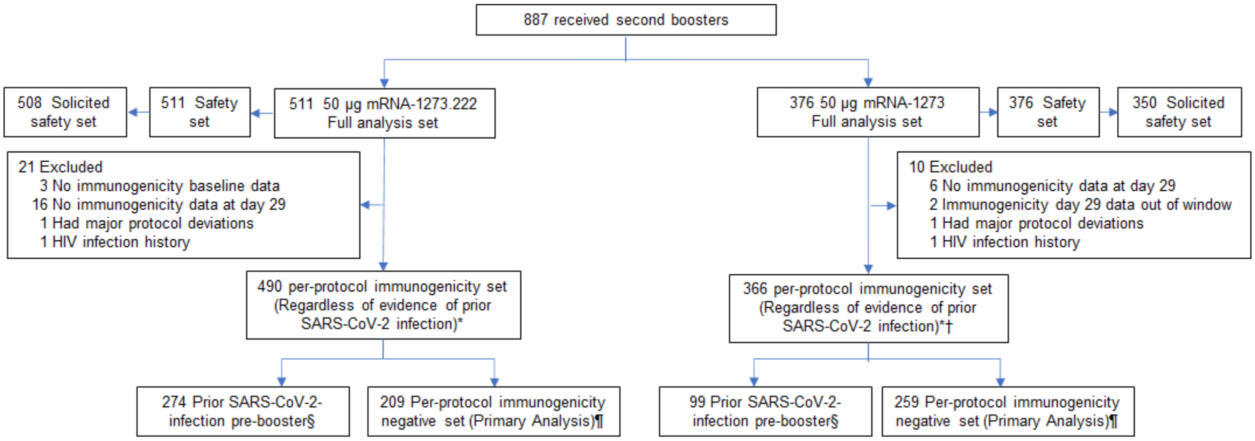


**Figure S1. Trial Profile of Analysis Sets.** The full analysis set consists of all participants who received study vaccine. The safety set consists of all participants who received study vaccine and was used for all analyses of safety except for solicited adverse reactions which were assessed in the solicited safety set. *The per-protocol set for immunogenicity consists of all participants in the full analysis set who received the planned dose of study vaccination and had antibody data available at pre-booster and day 29 and no major protocol deviations. †7 participants in the mRNA-1273.222 and 8 participants in the mRNA-1273 arm of the per-protocol immunogenicity set had missing pre-booster SARS-CoV-2 information. §Prior SARS-CoV-2-infection based on positive RT-PCR and/or serology test at baseline. ¶The per-protocol immunogenicity negative set (PPIS-negative) consists of participants in the per-protocol set for immunogenicity (PPIS) who have no serologic or virologic evidence of SARS-CoV-2 infection at baseline, i.e., who are SARS-CoV-2-infection negative, based on both negative RT-PCR tests for SARS-CoV-2 and negative SARS-CoV-2-nucleocapsid antibody test, and is the primary analysis set for immunogenicity analysis. A total of 379 participants received a second booster dose of 50-µg mRNA-1273; 2 participants had previously received the primary series but not a first booster dose, and another participant had a major protocol deviation. These three participants were excluded from all analysis sets. Data-cut-off date for mRNA-1273.222 was September 23, 2022.

**Figure S2: Statistical Testing Sequence for Immunogenicity Primary Objectives**

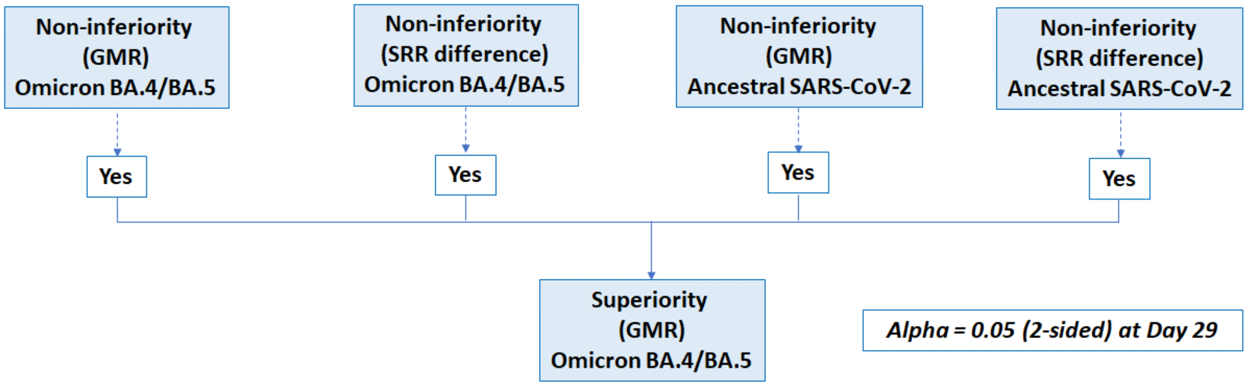


**Figure S2. Statistical Hypotheses Testing Strategy.**

For part H of the study, immunogenicity objectives were tested at day 29 with the family-wise type I error controlled at 0.05 (2-sided) At day 29, there were 4 corresponding clinical endpoints for the pre-specified primary objectives: non-inferiority of antibody response against omicron BA.4/BA.5 after the second booster doses of 50-µg mRNA-1273.222 versus 50-µg mRNA-1273 based on GMR (1) and SRR difference (2), and non-inferiority of the antibody response against SARS-COV-2 (D614G) after the second booster doses of 50-µg mRNA-1273.222 versus 50-µg mRNA-1273 based on GMR (3) and SRR difference (4) (detailed in Supplementary Statistical Methods). The endpoint testing sequence pre-specified that all 4 endpoints must first be met to test for the superiority of the antibody response against omicron BA.4/BA.5. All tests were based on alpha of 0.05 (2-sided).[9] Non-inferiority is considered met when the lower bound of the 95.0% confidence interval (CI) of GMR is >0.667 and the seroresponse rate-difference for BA.4/BA.5 is >-5% and seroresponse rate-difference for ancestral SARS-CoV-2 (D614G) is >-10%. Superiority is considered met when the lower bound of the 95.0% CI of GMR is >1.[5,6]

**Figure S3. Distribution of Observed Neutralizing Antibody Titers Against Omicron BA.4/BA.5 and SARS-CoV-2 (D614G) after 50-µg of mRNA-1273.222 and mRNA-1273 Administered as Second Booster Doses in Participants With and Without Prior SARS-CoV-2 Infection**

**A. Omicron BA.4/BA.5**

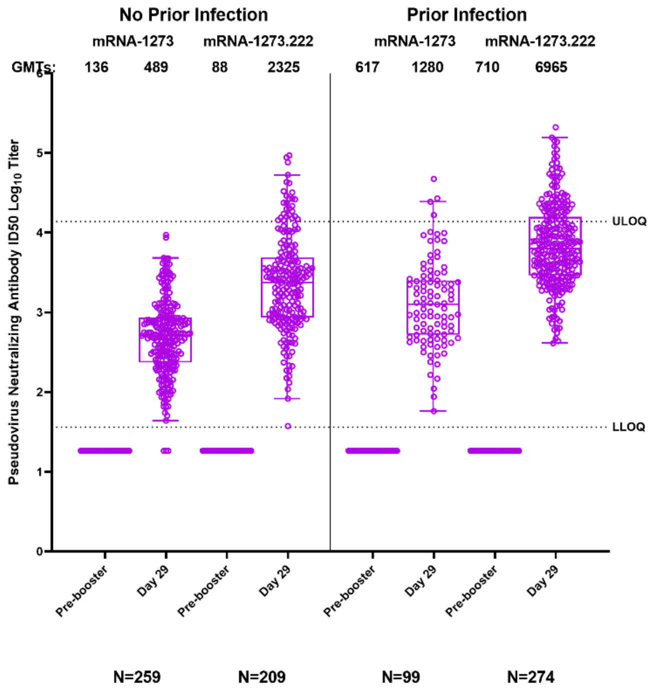


**B. Ancestral SARS-CoV-2 (D614G)**

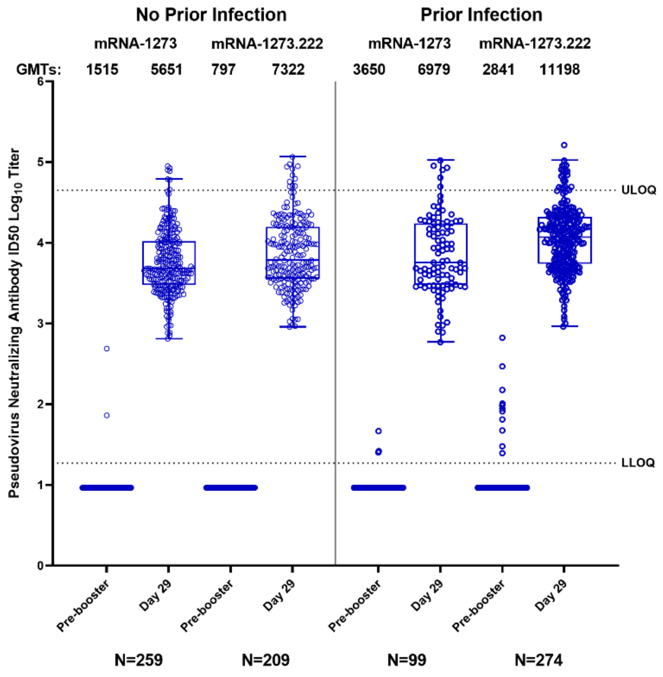

**Figure S3. Distribution of Observed Neutralizing Antibody Titers Against Omicron BA.4/BA.5 and SARS-CoV-2 (D614G) after 50-µg of mRNA-1273.222 and mRNA-1273 Administered as Second Booster Doses in Participants With and Without Prior SARS-CoV-2 Infection Doses.** Distribution of neutralizing antibody titers (ID50 $\log_{10}$) in the pseudovirus assay against omicron BA.4/BA.5 (Panel A) and ancestral SARS-CoV-2 (D614G) (Panel B) are shown for serum samples collected before the second booster dose of 50-µg of mRNA-1273.222 or 50-µg of mRNA-1273 (pre-booster), and at 28 days after the second booster dose (day 29) in those with and without prior SARS-CoV-2 infection. The circles are values from individual serum samples. Pseudovirus neutralizing antibody assay lower limits of quantification (LLOQ) are 18.5 (1.3 $\log_{10}$) for ancestral SARS-CoV-2 [D614G] and 36.7 (1.6 $\log_{10}$) for omicron BA.4/BA.5; upper limits of quantification (ULOQ) are 45,118 (4.7 $\log_{10}$) for ancestral SARS-CoV-2 [D614G] and 13,705 (4.1 $\log_{10}$) for omicron BA.4/BA.5. Boxes and horizontal bars denote interquartile (IQR) ranges and median endpoint titers; whisker endpoints are the maximum and minimum values below or above the median ±1.5 times the IQR. Refer to Table 2 for the observed geometric mean titers and geometric mean fold rises, and the estimated neutralizing antibody geometric mean titers and geometric mean ratios (ANCOVA model-based) following the second booster doses of 50-µg mRNA-1273 or 50-µg of mRNA-1273.214.

**Figure S4. Observed Neutralizing Antibody Response Omicron BA.4/BA.5 and Ancestral SARS-CoV-2 (D614G) after 50-µg of mRNA-1273.222 and mRNA-1273 Administered as Second Boosters in Participants with No Prior SARS-CoV-2-Infection by Age Groups**

### A. Omicron BA.4/BA.5

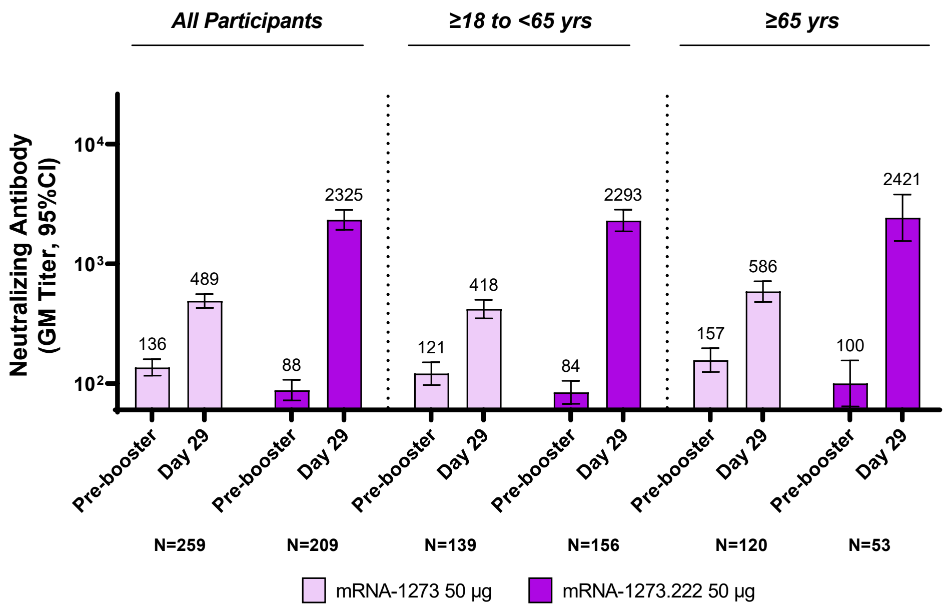


### B. Ancestral SARS-CoV-2 (D614G)

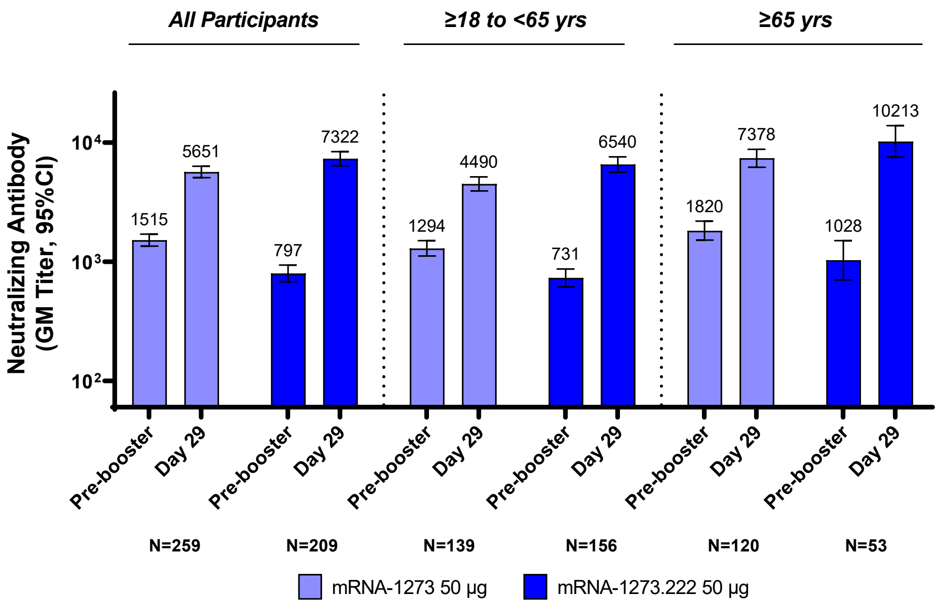

**Figure S5. Observed Neutralizing Antibody Titers Against Omicron Variants after 50-µg of mRNA-1273.222 and mRNA-1273.214 Administered as Second Booster Doses by Prior SARS-CoV-2 Infection Status at Pre-Booster**

**A. mRNA-1273.222**

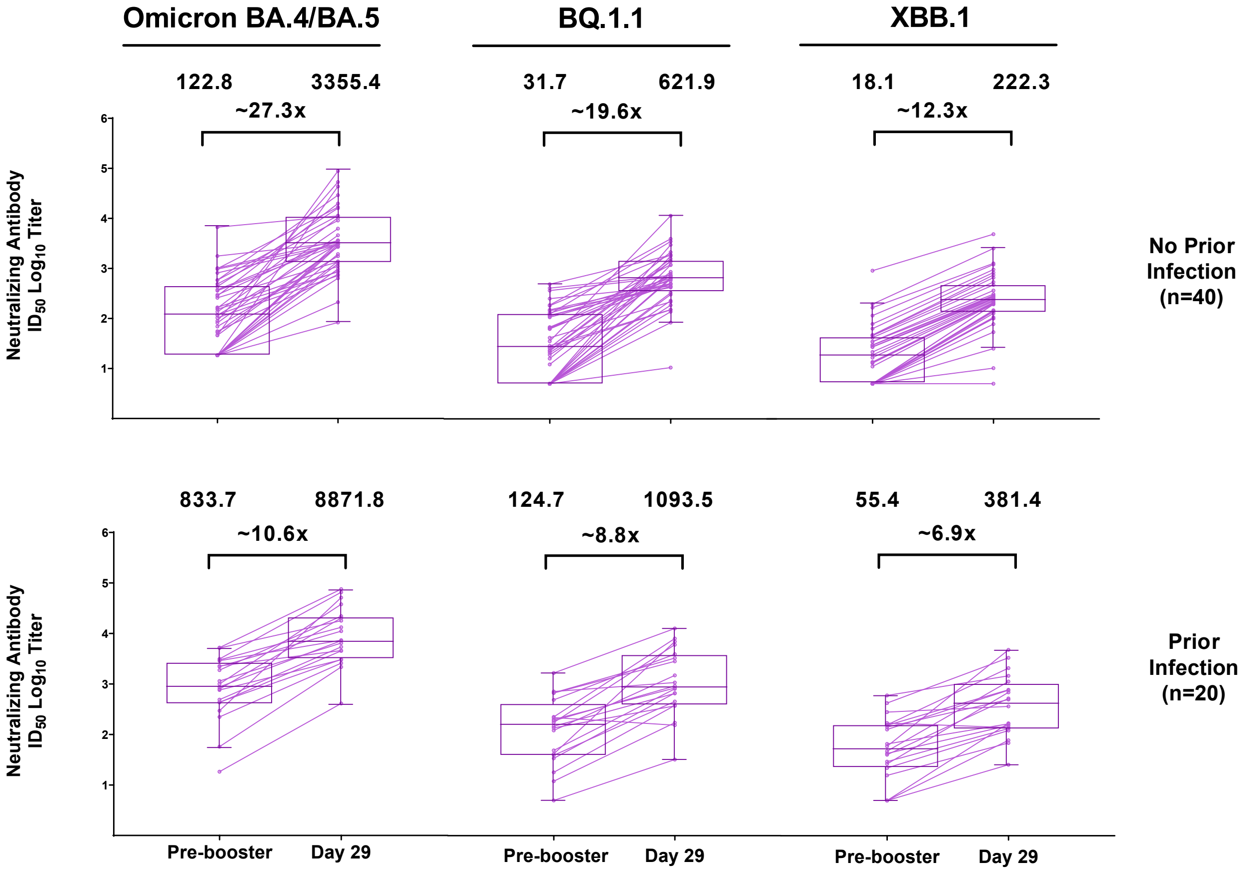

**B. mRNA-1273.214**

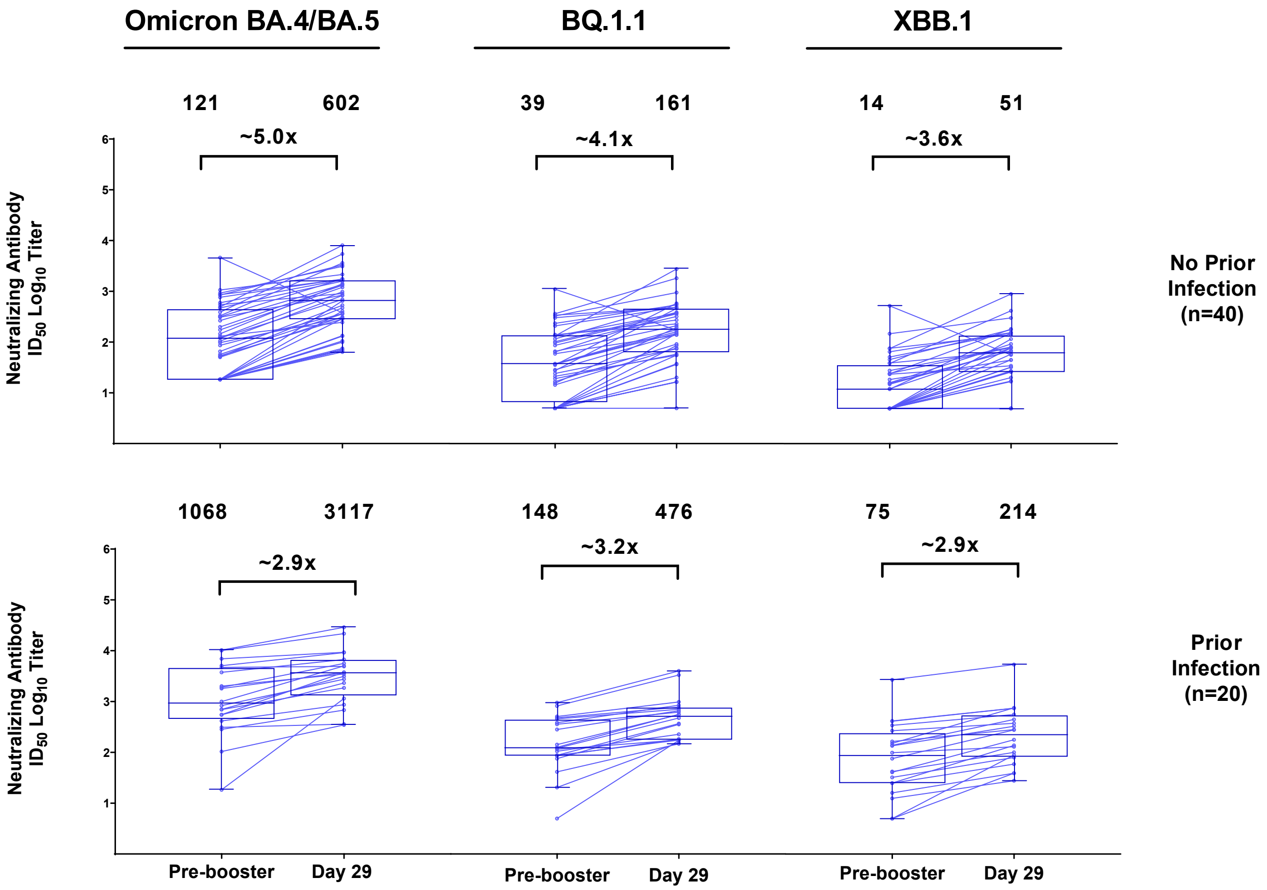


**Figure S5. Observed Neutralizing Antibody Titers Against Omicron Variants after 50-µg of mRNA-1273.222 and mRNA-1273.214 Administered as Second Booster Doses by Prior SARS-CoV-2 Infection Status at Pre-Booster mRNA-1273.222.** Pre-booster and day 29 neutralizing antibody titers ($log_{10}$) against omicron BA.4/BA.5, BQ1.1 and XBB.1 variants in the randomly-selected subsets of participants in the mRNA-1273.222 (panel A) and mRNA-1273.214 (panel B) booster groups, with (n=20) and without (n=40) prior SARS-CoV-2 infection. Antibody values reported as below the lower limit of quantification (LLOQ) are replaced by 0.5 x LLOQ. Values greater than the upper limit of quantification (ULOQ) are replaced by the ULOQ if actual values are not available. Omicron BA.4/BA.5 (ID50 LLOQ: 36.7, ULOQ: 13705). The limit of detection (LOD) of the PsVNA assay for BQ.1.1 and XBB.1 was 10; antibody values reported as below the LOD are replaced by 0.5 x LOD. Boxes and horizontal bars denote interquartile (IQR) ranges and median endpoint titers; whisker endpoints are the maximum and minimum values below or above the median ±1.5 times the IQR.

**Table S1. Objectives and Endpoints for Part H Second Booster Doses of 50-µg mRNA-1273.222 in Participants Who Received 100 µg mRNA-1273 Primary Series and a Booster Dose of 50 µg mRNA-1273**

| Objectives | Endpoints |
|---|---|
| **Primary** | |
| • To demonstrate non-inferiority of the antibody response of a second booster dose of mRNA-1273.222 50 µg compared to mRNA-1273 50 µg when administered as a second booster dose against Omicron BA.4/5 based on GMT ratio and SRR difference at Day 29<br>• To demonstrate superiority of the antibody response of a second booster dose of mRNA 1273.222 compared to mRNA-1273 (50 µg) administered as a second booster dose against the Omicron BA.4/5 based on GMT ratio at Day 29<br>• To demonstrate non-inferiority of the antibody response of mRNA-1273.222 50 µg compared to mRNA-1273 50 µg when administered as a second boosterdose against the ancestral SARS-CoV-2D614G based on GMT ratio and SRR difference at Day 29 | • GMT ratio of Omicron BA.4/5 GMT of mRNA-1273.222 over the Omicron BA.4/5 GMT of mRNA-1273 (Part F, Cohort 2, 50 µg mRNA-1273) at Day 29<br>• SRR difference between mRNA-1273.222 against Omicron BA.4/5 and mRNA-1273 against Omicron BA.4/5 at Day 29<br>• GMT ratio of the ancestral SARS-CoV-2 D614G GMT of mRNA-1273.222 over the ancestral SARS-CoV-2 D614G GMT of mRNA-1273 at Day 29<br>• SRR difference between mRNA-1273.222 and mRNA-1273 against the ancestral SARS-CoV-2 D614G at Day 29 |
| • To evaluate the safety and reactogenicity of mRNA 1273.222 | • Solicited local and systemic reactogenicity ARs during a 7-day follow-up period after vaccination<br>• Unsolicited AEs during the 28-day follow-up period after vaccination<br>• Serious AEs (SAEs), MAAEs, AEs leading to withdrawal and AESIs from Day 1 to end of study |
| **Secondary** | |
| To evaluate the immunogenicity of mRNA-1273.222 50 µg as a second booster dose against the ancestral SARS-CoV-2 (and other variants) compared to mRNA-1273 50 µg booster administered as a second booster dose at all timepoints post-boost | • GMT ratio of mRNA-1273.222 50 µg and mRNA-1273 50 µg against ancestral SARSCoV-2 (and other variants) at all timepoints post-booster<br>• SRR difference between mRNA-1273.222 50 µgand mRNA-1273 50 µg against ancestral SARSCoV-2 and variants of concern at all timepoints post-booster |
| **Exploratory** | |
| • To assess for symptomatic and asymptomatic SARS-CoV-2 infection | • Laboratory-confirmed symptomatic or asymptomatic SARS-CoV-2 infection will be defined in participants:<br>− Primary case definition per the (COVE) study<br>− Secondary case definition based on the CDC criteria: the presence of one of the CDC-listed symptoms (https://www.cdc.gov/coronavirus/2019-ncov/symptoms-testing/symptoms.html) and a |

| Objectives | Endpoints |
|---|---|
| | positive reverse transcriptase polymerase chain reaction (RT-PCR) test on a respiratory sample<br>• Asymptomatic SARS-CoV-2 infection is defined as a positive RT-PCR test on a respiratory sample in the absence of symptoms or a positive serologic test for anti-nucleocapsid antibody after a negative test at time of enrollment |
| • To evaluate the genetic and/or phenotypic relationships of isolated SARS-CoV-2 strains to the vaccine sequence | • Characterize the SARS-CoV-2 genomic sequence of viral isolates and compare with the vaccine sequence<br>• Characterize the immune responses to vaccine breakthrough isolates |

**Table S2. Analysis Sets**

| Set | Description |
| --- | --- |
| Full Analysis Set (FAS) | The FAS consists of all participants who receive investigational product (IP). |
| Per-Protocol (PP) Set for Immunogenicity | The PP Set for Immunogenicity consists of all participants in the FAS who received the planned dose of study vaccination and no major protocol deviations that impact key or critical data. |
| PP Set for Immunogenicity ‐ SARS-CoV-2 negative (PPSI-Neg) | Participants in the PPSI who have no serologic or virologic evidence of SARS-CoV-2 infection at baseline, i.e., who are SARS-CoV-2 negative, defined by both negative RT-PCR test for SARS-CoV-2 and negative serology test based on binding antibody (bAb) specific to SARS-CoV-2 nucleocapsid protein. PPSI-Neg will be the primary analysis set for analyses of immunogenicity for between booster comparisons. |
| Solicited Safety Set | The Solicited Safety Set consists of all participants who receive IP and contribute any solicited AR data.<br>The Solicited Safety Set will be used for the analyses of solicited ARs. Participants will be included in the study arm corresponding to the dose of IP that they actually received. |
| Safety Set | The Safety Set consists of all participants who receive IP. The Safety Set will be used for all analyses of safety except for the solicited ARs. Participants will be included in the study arm corresponding to the dose of IP that they actually received. |
| Per-Protocol Set for Efficacy | The PP Set for Efficacy consists of all participants in the FAS who receive the planned dose of study vaccination, who are SARS-CoV-2 negative at baseline (ie, have a negative RT-PCR test for SARS-CoV-2 and a negative serology test based on bAb specific to SARS-CoV-2 nucleocapsid at baseline), and have no major protocol deviations that impact key or critical data. |

### Table S3. Demographics for Subset of Participants Included in BQ1.1 and XBB.1 Variant Analysis

| Characteristics n (%)* | mRNA-1273.214 50 µg (N=60) | mRNA-1273.222 50 µg (N=60) |
|---|---|---|
| Age at Screening (yr) | | |
| Mean (range) | 57.7 (26-81) | 58.2 (25-82) |
| Age subgroup | | |
| ≥18 and <65 years | 30 (50.0) | 30 (50.0) |
| ≥65 years | 30 (50.0) | 30 (50.0) |
| Gender | | |
| Male | 21 (35.0) | 17 (28.3) |
| Female | 39 (65.0) | 43 (71.7) |
| Ethnicity | | |
| Hispanic or Latino | 8 (13.3) | 1 (1.7) |
| Not Hispanic or Latino | 52 (86.7) | 58 (96.7) |
| Not reported | 0 | 1 (1.7) |
| Race | | |
| White | 54 (90.0) | 55 (91.7) |
| Black or African American | 3 (5.0) | 4 (6.7) |
| Asian | 1 (1.7) | 1 (1.7) |
| American Indian or Alaska Native | 0 | 0 |
| Native Hawaiian or Other Pacific Islander | 0 | 0 |
| Multiracial | 1 (1.7) | 0 |
| Other | 1 (1.7) | 0 |
| Time between second injection of mRNA-1273 in primary series and first booster of mRNA-1273 (days) | | |
| n | 60 | 59 |
| Median | 247.5 | 245.0 |
| IQR | (223-278) | (228-324) |
| Time between first booster dose of mRNA-1273 and second booster (days) | | |
| n | 60 | 59 |
| Median | 135.0 | 293.0 |
| IQR | (116-146) | (266-318) |
| Pre-booster RT-PCR SARS-CoV-2 | | |
| Negative | 60 (100) | 56 (93.3) |
| Positive | 0 | 2 (3.3) |
| Missing | 0 | 2 (3.3) |
| Pre-booster antibody to SARS-CoV-2 nucleocapsid† | | |
| Negative | 40 (66.7) | 41 (68.3) |
| Positive | 20 (33.3) | 19 (31.7) |
| Missing | 0 | 0 |
| Pre-booster SARS-CoV-2 status‡ | | |
| Negative | 40 (66.7) | 40 (66.7) |
| Positive | 20 (33.3) | 20 (33.3) |
| Positive by both RT-PCR and SARS-CoV-2 nucleocapsid† | 0 | 1(1.7) |
| Positive by RT-PCR only | 0 | 1(1.7) |
| Positive by SARS-CoV-2 nucleocapsid only† | 20 (33.3) | 18 (30.0) |
| Missing | 0 | 0 |

IQR=interquartile range; RT-PCR = reverse transcription polymerase chain reaction. *Percentages based on the number of participants in the population set. †Elecsys assay for binding antibody to SARS-CoV-2 nucleocapsid. ‡Pre-booster SARS-CoV-2 status was positive if there was evidence of prior SARS-CoV-2-infection, defined as positive binding antibody against the SARS-CoV-2 nucleocapsid or positive RT-PCR at day 1; Negative SARS-CoV-2 status was defined as negative binding antibody against the SARS-CoV-2 nucleocapsid and a negative RT-PCR at day 1. Table - Demog

**Table S4. Solicited Local and Systemic Adverse Reactions Within 7 Days Following the Second Booster Injections of mRNA-1273.222 (50 µg) by Pre-booster SARS-CoV-2 Status, Solicited Safety Set**

| Adverse Reaction, N (%) | mRNA-1273.222 Second Booster Dose (50 µg) | | |
|---|---|---|---|
| | All participants N=508 | SARS-CoV-2 Negative N=216 | SARS-CoV-2 Positive N=283 |
| Solicited AR, N1 | 508 | 216 | 283 |
| Any Solicited AR | 443 (87.2) | 191 (88.4) | 244 (86.2) |
| Grade 1 | 219 (43.1) | 88 (40.7) | 129 (45.6) |
| Grade 2 | 165 (32.5) | 74 (34.3) | 87 (30.7) |
| Grade 3 | 59 (11.6) | 29 (13.4) | 28 (9.9) |
| Grade 4 | 0 | 0 | 0 |
| Any Solicited Local AR, N1 | 507 | 216 | 282 |
| Any Solicited Local AR | 420 (82.8) | 183 (84.7) | 229 (81.2) |
| Grade 1 | 329 (64.9) | 138 (63.9) | 187 (66.3) |
| Grade 2 | 63 (12.4) | 29 (13.4) | 32 (11.3) |
| Grade 3 | 28 (5.5) | 16 (7.4) | 10 (3.5) |
| Local AR, Pain, N1 | 507 | 216 | 282 |
| Pain | 418 (82.4) | 183 (84.7) | 227 (80.5) |
| Grade 1 | 344 (67.9) | 145 (67.1) | 194 (68.8) |
| Grade 2 | 54 (10.7) | 26 (12.0) | 26 (9.2) |
| Grade 3 | 20 (3.9) | 12 (5.6) | 7 (2.5) |
| Erythema, N1 | 507 | 216 | 282 |
| Erythema | 23 (4.5) | 11 (5.1) | 11 (3.9) |
| Grade 1 | 10 (2.0) | 5 (2.3) | 5 (1.8) |
| Grade 2 | 8 (1.6) | 4 (1.9) | 4 (1.4) |
| Grade 3 | 5 (1.0) | 2 (0.9) | 2 (0.7) |
| Swelling, N1 | 507 | 216 | 282 |
| Swelling | 40 (7.9) | 18 (8.3) | 21 (7.4) |
| Grade 1 | 22 (4.3) | 10 (4.6) | 12 (4.3) |
| Grade 2 | 13 (2.6) | 6 (2.8) | 7 (2.5) |
| Grade 3 | 5 (1.0) | 2 (0.9) | 2 (0.7) |
| Axillary Swelling or Tenderness, N1 | 507 | 216 | 282 |
| Axillary Swelling or Tenderness | 106 (20.9) | 46 (21.3) | 58 (20.6) |
| Grade 1 | 86 (17.0) | 37 (17.1) | 47 (16.7) |
| Grade 2 | 19 (3.7) | 8 (3.7) | 11 (3.9) |
| Grade 3 | 1 (0.2) | 1 (0.5) | 0 |
| Any Systemic AR, N1 | 508 | 216 | 283 |
| Any Systemic AR | 372 (73.2) | 167 (77.3) | 197 (69.6) |
| Grade 1 | 172 (33.9) | 76 (35.2) | 93 (32.9) |
| Grade 2 | 165 (32.5) | 75 (34.7) | 85 (30.0) |
| Grade 3 | 35 (6.9) | 16 (7.4) | 19 (6.7) |
| Fever, N1 | 507 | 216 | 282 |
| Fever | 20 (3.9) | 9 (4.2) | 10 (3.5) |

| | | | |
|---|---|---|---|
| Grade 1 | 12 (2.4) | 5 (2.3) | 6 (2.1) |
| Grade 2 | 7 (1.4) | 3 (1.4) | 4 (1.4) |
| Grade 3 | 1 (0.2) | 1 (0.5) | 0 |
| Headache, N1 | 507 | 216 | 282 |
| Headache | 249 (49.1) | 116 (53.7) | 126 (44.7) |
| Grade 1 | 164 (32.3) | 75 (34.7) | 84 (29.8) |
| Grade 2 | 73 (14.4) | 36 (16.7) | 35 (12.4) |
| Grade 3 | 12 (2.4) | 5 (2.3) | 7 (2.5) |
| Fatigue, N1 | 508 | 216 | 283 |
| Fatigue | 304 (59.8) | 144 (66.7) | 154 (54.4) |
| Grade 1 | 161 (31.7) | 77 (35.6) | 83 (29.3) |
| Grade 2 | 126 (24.8) | 57 (26.4) | 64 (22.6) |
| Grade 3 | 17 (3.3) | 10 (4.6) | 7 (2.5) |
| Myalgia, N1 | 507 | 216 | 282 |
| Myalgia | 235 (46.4) | 109 (50.5) | 121 (42.9) |
| Grade 1 | 127 (25.0) | 58 (26.9) | 66 (23.4) |
| Grade 2 | 88 (17.4) | 40 (18.5) | 46 (16.3) |
| Grade 3 | 20 (3.9) | 11 (5.1) | 9 (3.2) |
| Arthralgia, N1 | 507 | 216 | 282 |
| Arthralgia | 177 (34.9) | 78 (36.1) | 93 (33.0) |
| Grade 1 | 108 (21.3) | 48 (22.2) | 55 (19.5) |
| Grade 2 | 60 (11.8) | 24 (11.1) | 35 (12.4) |
| Grade 3 | 9 (1.8) | 6 (2.8) | 3 (1.1) |
| Nausea/ vomiting, N1 | 507 | 216 | 282 |
| Nausea / vomiting | 71 (14.0) | 31 (14.4) | 39 (13.8) |
| Grade 1 | 56 (11.0) | 26 (12.0) | 29 (10.3) |
| Grade 2 | 14 (2.8) | 5 (2.3) | 9 (3.2) |
| Grade 3 | 1 (0.2) | 0 | 1 (0.4) |
| Chills, N1 | 507 | 216 | 282 |
| Chills | 112 (22.1) | 51 (23.6) | 58 (20.6) |
| Grade 1 | 60 (11.8) | 27 (12.5) | 32 (11.3) |
| Grade 2 | 48 (9.5) | 23 (10.6) | 23 (8.2) |
| Grade 3 | 4 (0.8) | 1 (0.5) | 3 (1.1) |

AR=adverse reaction. Any=Grade 1 or higher (the highest grade for an event category was used in the summaries for participants reporting multiple events in the category). N1=Number of exposed participants with any information about the adverse event. Percentages are based on the number of participants who submitted any data for the event.

**Table S5. Summary of Unsolicited Adverse Events ≤28 Days Post-booster Dose and Up to Data Cutoff, Safety Set**

| | mRNA-1273.222 50 µg | |
|---|---|---|
| **n (%)** | **AEs ≤28 days post-booster** | **AEs up to data cutoff** |
| | **(N=511)** | |
| **Unsolicited AEs Regardless of Relationship to Study Vaccination** | | |
| All | 116 (22.7) | 129 (25.2) |
| Serious | 3 (0.6)* | 3 (0.6)* |
| Fatal | 1 (0.2)† | 1 (0.2)† |
| Medically attended | 70 (13.7) | 83 (16.2) |
| Leading to discontinuation from study | 0 | 0 |
| Grade 3 or higher | 5 (1.0) | 5 (1.0) |
| **Unsolicited AEs related to study vaccination** | | |
| All | 40 (7.8) | 40 (7.8) |
| Serious | 0 | 0 |
| Fatal | 0 | 0 |
| Medically attended | 0 | 0 |
| Leading to discontinuation from study | 0 | 0 |
| Grade 3 or higher | 2 (0.4)§ | 2 (0.4)§ |

AE=adverse event. An adverse event was defined as any event not present prior to study vaccination or any event already present that worsened in intensity or frequency after vaccination. Percentages are based on the number of participants in the safety set.

*Four serious AEs in three participants (60-70 years of age) were reported that were considered unrelated to study vaccination; these included events of anginal equivalent (chest pain, study day 4) and syncope (study day 10) and an event of anemia (study day 9) in two participants with medical history/risk factors for vascular and cardiac disease and of anemia requiring blood transfusions, gastro-oesophageal reflux disease, hepatic cirrhosis and positive pre-booster SARS-CoV-2 status, respectively, and †one fatal subarachnoid hemorrhage (study day 7) that occurred in a participant with a medical history of vascular and cardiac disease in the mRNA-1273.222 group.

§Two severe AEs of fatigue considered related to study vaccination occurred in the mRNA-1273.222 group which both started at day 2, one ended at day 8 and the other ended at day 28. One SAE with a fatal outcome due to an unknown cause occurred in the mRNA1273.222 group, considered unrelated to study vaccination after the data cutoff date (study day 40). Data-cut-off date was September 23, 2022.

### Table S6. Geometric Mean Neutralizing Antibody Titer Ratio and Seroresponse Difference Against Ancestral SARS-CoV-2 (D614G) and Omicron BA.4/BA.5 after 50-µg of mRNA-1273.222 and mRNA-1273 administered as Second Booster Doses in Participants with Evidence of SARS-CoV-2 Infection at Pre-booster

| | Ancestral SARS-CoV-2 (D614G) | | Omicron BA.4/BA.5 | |
|---|---|---|---|---|
| | 50 µg mRNA-1273.222 Booster Dose | 50 ug mRNA-1273 Booster dose | 50 µg mRNA-1273.222 Booster Dose | 50 ug mRNA-1273 Booster dose |
| | N=274 | N=99 | N=274 | N=99 |
| Pre-booster n† | 274 | 99 | 274 | 99 |
| Observed GMT (95% CI)§ | 2841.1 (2475.0, 3261.4) | 3649.5 (2758.5, 4828.2) | 710.2 (606.9, 831.1) | 616.8 (453.1, 839.8) |
| Day 29, n† | 274 | 99 | 274 | 99 |
| Observed GMT (95% CI)§ | 11197.9 (10035.1-12495.5) | 6979.3 (5585.6-8720.9) | 6964.5 (6043.7-8025.4) | 1280.2 (996.7-1644.3) |
| GMFR (95% CI)§ | 3.9 (3.5-4.4) | 1.9 (1.6-2.2) | 9.8 (8.4-11.4) | 2.1 (1.8-2.4) |
| Estimated GMT (95% CI)¶ | 12659.4 (11361.6-14105.4) | 6872.8 (5877.7-8036.2) | 7607.7 (6607.4-8759.5) | 1490.2 (1217.3-1824.4) |
| GMR (95.0% CI)¶ | 1.84 (1.56-2.18) | | 5.11 (4.10-6.36) | |
| Day 29 SRR injection 1, n/N1, %‖ (95% CI) | 174/174, 100 (97.9-100.0) | 79/79, 100 (95.4-100.0) | 162/162, 100 (97.7-100.0) | 74/76, 97.4 (90.8-99.7) |
| Difference (95.0% CI)‡ | 0 | | 5.5 (0.5-10.5) | |
| Day 29 SRR prebooster, n/N1, %‖ (95% CI) | 125/274, 45.6 (39.6-51.7) | 17/99, 17.2 (10.3-26.1) | 203/274, 74.1 (68.5-79.2) | 17/99, 17.2 (10.3-26.1) |
| Difference (95.0% CI)‡ | 27.6 (18.2-37.1) | | 55.3 (46.2-64.4) | |

ANCOVA=analysis of covariance, CI=confidence interval, GMT=geometric mean titer, GMFR= geometric mean fold rise (post-baseline/baseline titers), GMR=geometric mean ratio (mRNA-1273.222 vs mRNA-1273). ID50=50% inhibitory dose; LLOQ=lower limit of quantification; LS=least squares; SRR=seroresponse rate; ULOQ=upper limit of quantification. Antibody values assessed by pseudovirus neutralizing antibody assay reported as below the LLOQ (18.5 for ancestral [D614G] and 36.7 for omicron BA.4/BA.5 are replaced by 0.5 × LLOQ. Values greater than ULOQ (45,118) for ancestral [D614G] and (13,705) for omicron BA.4/BA.5 are replaced by the ULOQ if actual values are not available.

N1= Number of participants with non-missing data at baseline (injection 1/pre-booster) and the corresponding post-baseline timepoint.

†Number of participants with non-missing data at the timepoint (baseline or post-baseline).

§95% CI based on the t-distribution of log-transformed values or difference in the log-transformed values for GMT value and GMFR, respectively, then back transformed to the original scale.

¶Log-transformed antibody levels are analyzed using an ANCOVA model with the treatment variable as fixed effect, adjusting for age group (<65, ≥65 years) and pre-booster titers. The resulting LS means, difference of LS means, and 95% CI are back transformed to the original scale for presentation.

‖Seroresponse at a participant level defined as a change from <LLOQ to ≥4 × LLOQ, or at least a 4-fold rise if baseline is ≥LLOQ; comparison to pre-vaccination/pre-booster baselines. Percentages were based on the number of participants with non-missing data at baseline and the corresponding time point. 95% CI calculated using the Clopper-Pearson method.

‡The SRR difference is a calculated common risk difference adjusted by age group using inverse-variance stratum weights and the middle point of Miettinen-Nurminen confidence limits of each one of the stratum risk differences. The stratified Miettinen-Nurminen estimate and the CI cannot be calculated when the seroresponse rate in both groups is 100%, absolute difference is reported.

**Table S7. Neutralizing Omicron BA.4/BA.5 Neutralizing Titers by Time Intervals Between First Booster Dose of mRNA-1273 and Second Booster Doses of mRNA-1273.222 and mRNA-1273 in Participants Without Prior Infection**

| | Quartile dosing time intervals (days) between first booster dose of mRNA-1273 and second boosters | | | |
|---|---|---|---|---|
| **mRNA-1273.222** | **<258 (N=47)** | **258 - <289 (N=58)** | **289 - <312 days (N=49)** | **≥312 (N=55)** |
| Pre-booster | | | | |
| n* | 47 | 58 | 49 | 55 |
| GMT Level | 113.7 | 87.9 | 73.3 | 82.9 |
| 95% CI | (76.9-168.2) | (63.0-122.8) | (47.8-112.6) | (52.8-130.3) |
| Day 29 | | | | |
| n* | 47 | 58 | 49 | 55 |
| GM Level | 2726.5 | 3517.9 | 1697.2 | 1734.3 |
| 95% CI† | (1775.2-4187.7) | (2475.1-5000.1) | (1120.5-2570.8) | (1235.2-2435.1) |
| GMFR | 24.0 | 40.0 | 23.1 | 20.9 |
| 95% CI† | (16.6-34.5) | (27.8-57.6) | (15.5-34.6) | (14.5-30.1) |
| **mRNA-1273** | **<118 (N=65)** | **118 days - <134 (N=71)** | **134 - <150 (N=59)** | **≥150 (N=63)** |
| Pre-booster | | | | |
| n* | 65 | 71 | 59 | 63 |
| GMT Level | 137.7 | 177.2 | 139.7 | 98.0 |
| 95% CI | (101.0-187.8) | (130.8-239.9) | (100.7-193.9) | (70.4-136.3) |
| Day 29 | | | | |
| n* | 65 | 71 | 59 | 63 |
| GM Level | 423.1 | 561.8 | 514.8 | 464.0 |
| 95% CI† | (319.3, 560.6) | (438.7, 719.3) | (404.3, 655.4) | (339.1, 635.0) |
| GMFR | 3.1 | 3.2 | 3.7 | 4.7 |
| 95% CI† | (2.5-3.7) | (2.6-3.8) | (3.0-4.5) | (3.9-5.7) |

CI=confidence interval, GM=geometric mean, GMFR=geometric mean fold-rise (post-baseline/baseline titers). Antibody values reported as below the lower limit of quantification (LLOQ) are replaced by 0.5 x LLOQ. Values greater than the upper limit of quantification (ULOQ) are replaced by the ULOQ if actual values are not available., Omicron BA.4/BA.5 (ID50 LLOQ: 36.7, ULOQ: 13705).
n= Number of participants with non-missing data at the timepoint (baseline or post-baseline).
†95% CI is calculated based on the t-distribution of the log-transformed values or the difference in the log-transformed values for GM value and GM fold-rise, respectively, then back transformed to the original scale for presentation.

**Table S8. Observed Binding Antibody Titers and Geometric Mean Ratios Against Ancestral SARS-CoV-2 (D614G) and Variants after 50-µg of mRNA-1273.222 and mRNA-1273 Administered as Second Booster Doses in Participants with No Evidence of SARS-CoV-2 Infection at Pre-Booster**

**A. Ancestral (D614G) and Omicron BA.1**

| | Ancestral SARS-CoV-2 (D614G) | | Omicron BA.1 | |
|---|---|---|---|---|
| | mRNA-1273.222 50 µg N=209 | mRNA-1273 50 µg N=259 | mRNA-1273.222 50 µg N=334 | mRNA-1273 50 µg N=259 |
| Pre-booster, n† | 209 | 259 | 209 | 259 |
| Observed GM levels§ | 120499.9 (105390.7-137775.3) | 279483.7 (254946.7-306382.2) | 25609.7 (22161.8-29593.9) | 56564.1 (51321.2-62342.6) |
| Day 29, n† | 208 | 259 | 208 | 259 |
| Observed GM Level (95% CI)§ | 845092.4 (764002.5-934789.2) | 796220.3 (726144.6-873058.6) | 228114.0 (206237.4-252311.1) | 177980.6 (162188.7-195310.2) |
| GMFR (95% CI)§ | 7.0 (6.2-7.8) | 2.8 (2.7-3.0) | 8.9 (7.9-10.1) | 3.1 (2.9-3.4) |
| Estimated GM Level (95% CI)¶ | 1144974.8 (1053713.8-1244139.7) | 653278.1 (608076.3, 701840.1) | 296366.9 (272274.2-322591.4) | 149765.9 (139239.1-161088.6) |
| GMR (95% CI)¶ | 1.75 (1.57-1.96) | | 1.98 (1.77-2.22) | |

**B. Other variants**

| | Alpha | | Gamma | | Delta | |
|---|---|---|---|---|---|---|
| | mRNA-1273.222 50 µg N=209 | mRNA-1273 50 µg N=259 | mRNA-1273.222 50 µg N=209 | mRNA-1273 50 µg N=259 | mRNA-1273.222 50 µg N=209 | mRNA-1273 50 µg N=259 |
| Pre-booster, n† | 209 | 259 | 209 | 259 | 209 | 259 |
| Observed GM levels§ | 93589.5 (81491.1-107484.1) | 206919.5 (188334.0-227339.1) | 63411.1 (55197.0-72847.6) | 135931.8 (123850.4-149191.8) | 90420.0 (78985.0-103510.5) | 175659.4 (159935.1-192929.6) |
| Day 29, n† | 208 | 259 | 208 | 259 | 208 | 259 |
| Observed GM Level (95% CI)§ | 665136.7 (600686.7-736501.6) | 598907.1 (546449.4-656400.6) | 516162.8 (465447.0-572404.7) | 396861.4 (362419.4-434576.4) | 652079.7 (590834.9-719673.1) | 519695.1 (476509.6-566794.4) |
| GMFR (95% CI)§ | 7.1(6.3-8.0) | 2.9 (2.7-3.1) | 8.1 (7.2-9.1) | 2.9 (2.7-3.1) | 7.2 (6.4-8.1) | 3.0 (2.8-3.1) |
| Estimated GM Level (95% CI)¶ | 883192.7 (813896.6-958388.7) | 497096.4 (463311.9-533344.4) | 674537.4 (620463.9-733323.5) | 333070.1 (309979.6-357880.7) | 823169.0 (761676.1-889626.5) | 447514.2 (418579.9-478448.6) |
| GMR (95% CI)¶ | 1.78 (1.59-1.99) | | 2.03 (1.81- 2.27) | | 1.84 (1.66-2.04) | |

CI=confidence interval, GM=geometric mean, GMFR=geometric mean fold-rise (post-baseline/baseline titers), GMR=geometric mean ratio (mRNA-1273.222 relative to mRNA-1273), LLOQ=lower limit of quantification; Meso Scale Discovery=MSD; ULOQ=upper limit of quantification. Binding antibody values assessed by MSD assay reported as below the LLOQ (69 for ancestral SARS-CoV-2; 102 for omicron BA.1; 52 for alpha; 143 for gamma; 150 for delta) were replaced by 0.5 × LLOQ. Values greater than ULOQ (14,400,000 for ancestral SARS-CoV-2; 1,180,000 for omicron BA.1; 8,800,000 for alpha; 5,800,000 for gamma; 8,000,000 for delta) were replaced by the ULOQ if actual values are not available. Participant immune response data is censored at the last date of study participation (study discontinuation, study completion, or death), non-study COVID-19 vaccination date, or data cutoff/extraction date, whichever is the earliest.

†Number of participants with non-missing data at the timepoint (baseline or post-baseline). §95% CI based on the t-distribution of log-transformed values or difference in the log-transformed values for GMT value and GMT fold-rise, respectively, then back transformed to the original scale. ¶Log-transformed antibody levels are analyzed using an ANCOVA model with the treatment variable as fixed effect, adjusting for age group (<65, ≥65 years) and pre-booster titers. The resulting LS means, difference of LS means, and 95% CI are back transformed to the original scale for presentation.

### Table S9. Observed Binding Antibody Levels and Geometric Mean Ratios Against Ancestral SARS-CoV-2 (D614G) and Variants After Second 50-µg Boosters of mRNA-1273.222 and mRNA-1273 in Participants with Evidence of SARS-CoV-2 Infection at Pre-booster

#### A. Ancestral (D614G) and Omicron BA.1

| | Ancestral SARS-CoV-2 (D614G) | | Omicron BA.1 | |
|---|---|---|---|---|
| | mRNA-1273.222 50 µg N=274 | mRNA-1273 50 µg N=99 | mRNA-1273.222 50 µg N=274 | mRNA-1273 50 µg N=99 |
| Pre-booster, n† | 274 | 99 | 274 | 99 |
| Observed GM levels§ | 258113.8 (229970.7-289701.0) | 358525.8 (290554.2-442398.4) | 52662.3 (46348.6-59836.2) | 69048.6 (54420.3-87609.2) |
| Day 29, n† | 274 | 99 | 274 | 99 |
| Observed GM Level (95% CI)§ | 857657.7 (783912.2-938340.6) | 695725.8 (580841.4-833333.3) | 223117.3 (201877.4-246591.9) | 148377.3 (121556.3-181116.3) |
| GMFR (95% CI)§ | 3.3 (3.1-3.6) | 1.9 (1.8-2.1) | 4.2 (3.9-4.6) | 2.1 (2.0-2.4) |
| Estimated GM Level (95% CI)¶ | 978005.6 (907698.6-1053758.4) | 645146.8 (578653.2-719281.2) | 248115.4 (229130.3-268673.6) | 138318.9 (123259.1-155218.7) |
| GMR (95% CI)¶ | 1.52 (1.35-1.71) | | 1.79 (1.58- 2.03) | |

#### B. Other variants

| | Alpha | | Gamma | | Delta | |
|---|---|---|---|---|---|---|
| | mRNA-1273.222 50 µg N=274 | mRNA-1273 50 µg N=99 | mRNA-1273.222 50 µg N=274 | mRNA-1273 50 µg N=99 | mRNA-1273.222 50 µg N=274 | mRNA-1273 50 µg N=99 |
| Pre-booster, n† | 274 | 99 | 274 | 99 | 274 | 99 |
| Observed GM levels§ | 214881.7 (191118.0-241600.3) | 281608.0 (226599.9-349969.5) | 155707.8 (138998.9-174425.2) | 196151.1 (161524.5-238200.7) | 205035.6 (183477.3-229126.9) | 255142.6 (209140.3-311263.7) |
| Day 29, n† | 274 | 99 | 274 | 99 | 274 | 99 |
| Observed GM Level (95% CI)§ | 708888.9 (647227.7-776424.6) | 549296.6 (457295.2-659807.5) | 595259.4 (544534.4-650709.5) | 396864.7 (336237.0-468424.2) | 706974.9 (649218.8-769869.1) | 505719.7 (429970.1-594814.4) |
| GMFR (95% CI)§ | 3.3 (3.0-3.6) | 2.0 (1.8-2.1) | 3.8 (3.5-4.2) | 2.0 (1.9-2.2) | 3.4 (3.2-3.7) | 2.0 (1.8-2.2) |
| Estimated GM Level (95% CI)¶ | 796415.3 (739031.9-858254.4) | 519614.8 (466171.6-579184.8) | 666347.5 (617049.9-719583.5) | 385819.1 (345130.8-431304.2) | 791520.2 (735908.6, 851334.2) | 498215.6 (448166.5, 553854.0) |
| GMR (95% CI)¶ | 1.53 (1.36-1.72) | | 1.73 (1.53-1.95) | | 1.59 (1.42-1.78) | |

CI=confidence interval, GM=geometric mean, GMFR=geometric mean fold-rise (post-baseline/baseline titers), GMR=geometric mean ratio (mRNA-1273.222 relative to mRNA-1273), LLOQ=lower limit of quantification; Meso Scale Discovery=MSD; ULOQ=upper limit of quantification. Binding antibody values assessed by MSD assay reported as below the LLOQ (69 for ancestral SARS-CoV-2; 102 for omicron BA.1; 52 for alpha; 143 for gamma; 150 for delta) were replaced by 0.5 × LLOQ. Values greater than ULOQ (14,400,000 for ancestral SARS-CoV-2; 1,180,000 for omicron BA.1; 8,800,000 for alpha; 5,800,000 for gamma; 8,000,000 for delta) were replaced by the ULOQ if actual values are not available. Participant immune response data is censored at the last date of study participation (study discontinuation, study completion, or death), non-study COVID-19 vaccination date, or data cutoff/extraction date, whichever is the earliest.

†Number of participants with non-missing data at the timepoint (baseline or post-baseline). §95% CI based on the t-distribution of log-transformed values or difference in the log-transformed values for GMT value and GMT fold-rise, respectively, then back transformed to the original scale. ¶Log-transformed antibody levels are analyzed using an ANCOVA model with the treatment variable as fixed effect, adjusting for age group (<65, ≥65 years) and pre-booster titers. The resulting LS means, difference of LS means, and 95% CI are back transformed to the original scale for presentation.

**Table S10. Observed Neutralizing Antibody Geometric Mean Titers Against Omicron Variants BA.4/BA.5, BQ1.1 and XBB.1 after 50-µg of mRNA-1273.222 and mRNA-1273.214 Administered as Second Booster Doses by Prior SARS-CoV-2 Infection Status at Pre-Booster**

| | BA.4/BA.5 | | BQ.1.1 | | XBB.1 | |
|---|---|---|---|---|---|---|
| **Participants without Prior SARS-CoV-2 Infection** | **mRNA-1273.222 50 µg N=40** | **mRNA-1273.214 50 µg N=40** | **mRNA-1273.222 50 µg N=40** | **mRNA-1273.214 50 µg N=40** | **mRNA-1273.222 50 µg N=40** | **mRNA-1273.214 50 µg N=40** |
| Pre-booster, n* | 40 | 40 | 40 | 40 | 40 | 36 |
| GMT, 95% CI† | 122.8 (74.3-203.1) | 121.1 (74.6-196.4) | 31.7 (19.6-51.3) | 39.2 (23.9-64.3) | 18.1 (12.0-27.1) | 14.4 (9.5-21.7) |
| Day 29, n* | 40 | 40 | 40 | 40 | 40 | 39 |
| Observed GMTs (95% CI) † | 3355.4 (2109.9-5336.2) | 602.1 (399.9-906.4) | 621.9 (422.2-916.2) | 161.1 (104.1-249.3) | 222.3 (147.4-335.2) | 50.6 (32.4-79.2) |
| GMFR (95% CI)† | 27.3 (15.9-47.0) | 5.0 (3.7-6.7) | 19.6 (11.7-32.8) | 4.1 (3.0-5.5) | 12.3 (7.4-20.5) | 3.6 (2.5-5.1) |
| Fold-change vs BA.4/BA.5 GMT | 1 | 1 | 5.4 | 3.7 | 15.1 | 11.9 |
| **Participants with Prior SARS-CoV-2 Infection** | **N=20** | **N=20** | **N=20** | **N=20** | **N=20** | **N=20** |
| Pre-booster, n* | 20 | 20 | 20 | 20 | 20 | 20 |
| GMT (95% CI)† | 833.7 (422.5-1645.1) | 1067.6 (506.2-2251.7) | 124.7 (61.4-253.2) | 147.8 (80.9-270.0) | 55.4 (28.4-108.0) | 75.1 (35.4-159.1) |
| Day 29 | 20 | 20 | 20 | 20 | 20 | 20 |
| Observed GMTs (95% CI) | 8871.8 (4809.7-16364.8) | 3116.8 (1776.8-5467.3) | 1093.5 (536.8-2227.9) | 475.5 (304.7-742.0) | 381.4 (198.1-734.4) | 214.2 (116.9-392.4) |
| GMFR (95% CI) | 10.6 (6.4-17.6) | 2.9 (1.9-4.4) | 8.8 (5.0-15.5) | 3.2 (2.3-4.5) | 6.9 (4.0-11.7) | 2.9 (2.1-3.9) |
| Fold-change vs BA.4/BA.5 GMT | 1 | 1 | 8.1 | 6.6 | 23.3 | 14.6 |

CI=confidence interval, GM=geometric mean, GMFR=geometric mean fold rise (post-baseline / baseline titers).
N1=Number of subjects with non-missing data at baseline and the corresponding post-baseline timepoint.
Antibody values reported as below the lower limit of quantification (LLOQ) are replaced by 0.5 x LLOQ. Values greater than the upper limit of quantification (ULOQ) are replaced by the ULOQ if actual values are not available. Omicron BA.4/BA.5 (ID50: LLOQ:36.7, ULOQ:13705). The limit of detection (LOD) of the PsVNA assay for BQ.1.1 and XBB.1 was 10; antibody values reported as below the LOD are replaced by 0.5 x LOD.
*Number of participants with non-missing data at the timepoint (baseline or post-baseline).
†95% CI is calculated based on the t-distribution of the log-transformed values or the difference in the log-transformed values for GM value and GM fold-rise, respectively, then back transformed to the original scale for presentation.

### Table S11. Incidences of SARS-CoV-2-infection and Covid-19 Starting 14 Days Post-booster

| n (%) | Full Analysis Set<br>All Participants |
|---|---|
| | mRNA-1273.222 50 µg<br>N=511 |
| Covid-19 (COVE definition)*<br>Cases | 6 (1.2) |
| Covid-19 (CDC definition)*<br>Cases | 8 (1.6) |
| SARS-CoV-2 infection (regardless of symptoms)<br>Cases | 17 (3.3) |
| Asymptomatic SARS-CoV-2 infection<br>Cases | 9 (1.8)† |

*Covid-19 case definitions are based on 2 systemic symptoms or at least one of the respiratory symptoms used in the Coronavirus Efficacy (COVE) trial [1,2] and Covid-19 cases based on 1 symptom per CDC definition.
The full analysis set consists of all participants who received study vaccine. Covid-19, SARS-CoV-2 infection, and asymptomatic SARS-CoV-2 infection are censored at the earlier date of last date of study participation, non-study COVID-19 vaccination date, or efficacy data cutoff/extraction date.
†Of these 9 cases, 5 cases were identified by PCR and 4 were identified by serology tests (anti-nucleocapsid assay).
Data-cut-off date for mRNA-1273.222 was September 23, 2022.
